# Visionary AI: Decoding Systemic Vascular Health and Hypertensive Disorders in Pregnancy Through Retinal Imaging and Artificial Intelligence

**DOI:** 10.1101/2025.11.25.25340974

**Authors:** Srilaxmi Bearelly, Cassandra Areff, Matthew K. Hoffman, Daniel Sunko, Andrew F. Laine, Bin Choi, Hanna Rodriguez Coleman, Bianca Cordazzo Vargas, Chloe Y. Li, Jennifer Dawkins, Emily Amir, Lisa A. Hark, Whitney A. Booker, April Jauhal, Ronald J. Wapner, Liat Shenhav

## Abstract

Pregnancy orchestrates a rare physiological transformation across vascular, immune, and metabolic systems. When this dynamic balance is disrupted – as in hypertensive disorders of pregnancy – the consequences can be life-threatening, spanning maternal mortality, fetal growth restriction, and elevated long-term cardiovascular risk. Despite clear links to early placental dysfunction and systemic endothelial disruption, current screening remains clinically imprecise, biologically opaque, and logistically challenging. A shift is urgently needed – from detecting maternal complications late in gestation to understanding how pregnancy reshapes vascular physiology systemically and how this remodeling may go awry.

Here we present Visionary AI, an artificial intelligence platform that integrates ultra-widefield retinal imaging (200°) with biologically grounded vascular modeling to predict hypertensive disorders of pregnancy early in gestation. Unlike prior approaches that rely on generic deep learning models and clinical inputs, Visionary AI constructs an interpretable, graph-based representation of the maternal retinal vasculature and applies topological and geometric analysis to identify condition-specific microvascular signatures. In a prospective multiethnic U.S. cohort of 1,267 pregnancies, Visionary AI achieved high predictive performance for preeclampsia (AUC = 0.90), early-onset (AUC = 0.93), and severe preeclampsia (AUC = 0.89), outperforming current clinical paradigms. It also generalized to predict gestational hypertension (AUC = 0.91) and chronic hypertension (AUC = 0.90).

Topological and geometric analyses of the vasculature revealed distinct and interpretable remodeling patterns across subtypes of hypertensive disorders of pregnancy, offering mechanistic insight into their divergent pathophysiology.

These results position the maternal retina as a minimally invasive, high-fidelity biosensor of early systemic vascular health and establish Visionary AI as a clinically actionable, biologically grounded diagnostic framework with potential for broad global scalability.

## Introduction

### Hypertensive Disorders of Pregnancy

Hypertensive disorders of pregnancy (HDP), including gestational hypertension (GHTN), preeclampsia, chronic hypertension with superimposed preeclampsia, and eclampsia, affect 2– 15% of pregnancies and represent the second leading cause of maternal mortality worldwide^1^. HDPs increase the risk of acute maternal complications such as renal dysfunction, pulmonary edema, stroke, and death^2^, and contribute to adverse fetal outcomes, including small-for-gestational-age, preterm delivery, and stillbirth^3^. Long-term, they are associated with elevated maternal risk of hypertension, coronary artery disease, stroke, and vascular dementia^4–6^, and elevated offspring risk of cardiovascular and neurodevelopmental disorders^7^.

Despite their burden, HDPs are typically diagnosed after 20 weeks’ gestation, when symptoms such as hypertension, proteinuria, or organ dysfunction become clinically apparent. Severe features – including blood pressure (BP) >160/110 mmHg, thrombocytopenia, and visual symptoms – guide early delivery decisions due to high maternal–fetal risk^8^. Early-onset preeclampsia (EOPE, symptoms appear before 34 weeks) tends to present more severe complications than late-onset preeclampsia (LOPE). Notably, recent systems biology studies suggest that preeclampsia is not a single disease entity but may reflect multiple distinct biological pathways – with EOPE more frequently associated to placental dysfunction and LOPE more often associated with maternal processes^9,10^.

### Pathophysiology and Prevention

Mechanistically, preeclampsia is believed to arise, in part, from maternal vascular inflammation and thrombosis that disrupt placental development^9,11^. Inflammation and thrombosis are known to inhibit deep placental invasion^12^ of the maternal decidua between 10 and 16 weeks’ gestation and then further impair the placenta resulting in placental atherosis^13,14,15,16^, a process very similar to cardiovascular disease. These early deviations from physiologic placentation precede clinical symptoms by weeks or months, underscoring the need for early biomarkers of systemic and placental vascular stress.

From a health systems perspective, HDPs accounted for an estimated $2.18 billion in U.S. healthcare costs per year (estimated in 2012), largely driven by preterm birth complications^17^. Their incidence has increased in subsequent years^18,19,20^, and pregnancies complicated by HDPs typically end ∼3 weeks earlier than normotensive pregnancies^21^. Importantly, randomized trials show that early initiation of low-dose aspirin (before 16 weeks) can reduce risk of preterm preeclampsia by up to 62%^22,23^ – but this requires early, accurate risk identification.

### Limitations of Current Screening

Current diagnostic criteria for HDPs rely on late-onset symptoms (hypertension, proteinuria, end-organ dysfunction), usually manifesting in the third trimester^24^. While known risk factors (e.g., prior HDPs, diabetes, obesity, age) provide some predictive value, they lack individual-level accuracy^25^. The sFlt-1/PlGF ratio is the most validated biomarker but is only useful after 20 weeks and in symptomatic patients^26^. First-trimester screening tools, such as the Fetal Medicine Foundation calculator^27^, combine clinical variables with biomarkers or ultrasound, but these protocols are expensive, logistically complex, and inconsistently implemented^28,29^.

Taken together, these gaps underscore the need for a non-invasive, cost-effective, and scalable screening tool that enables accurate risk prediction in the first trimester, prior to symptom onset, and is suitable for broad population-level deployment.

### Retinal Vascular Biomarkers of Hypertensive Disorders of Pregnancy

Increasing evidence suggests a link between placental vascular changes associated with preeclampsia and alterations in the retinal vasculature. Studies of women with established preeclampsia describe retinal signs resembling hypertensive retinopathy, arteriolar narrowing, and choroidal changes^30–34^, and link preeclampsia to increased future risk of retinal diseases^35^. Further, recent evidence has identified an association between the sFlt-1/PlGF ratio and choroidal thickness as measured by optical coherence tomography (OCT)^33,36^. A prior postpartum study from our team (31 severe preeclampsia cases and 35 normotensive controls) demonstrated significantly increased median tortuosity in the inferotemporal retinal artery among preeclampsia patients^37^. OCT angiography (OCT-A) has revealed that pregnant women who go on to develop placental insufficiency have significantly narrower retinal blood vessels (arterioles and venules) compared to normotensive pregnancies^38^. These patients also exhibit reduced capillary density. In separate studies, women with preeclampsia had significantly lower vessel densities in both the superficial and deep capillary plexuses – including in the foveal and parafoveal regions – relative to healthy pregnant or non-pregnant controls^39,40^. These findings suggest a degree of capillary dropout or “vascular rarefaction” in the retinas of women with preeclampsia. Notably, such changes are often subclinical. Retinal vessel caliber reduction and reduced perfusion density likely result from systemic vasoconstriction and endothelial dysfunction, both hallmarks of preeclampsia pathophysiology. Importantly, these changes may be subclinical and observable even in early pregnancy, mirroring processes of endothelial dysfunction and systemic vasoconstriction.

### AI-Powered Retinal Imaging for Pregnancy Risk Stratification

Retinal imaging offers a scalable, non-invasive method for systemic vascular phenotyping. Ultra-widefield (UWF) systems can capture ∼200° of the retina in under 2 minutes without pupil dilation, making them highly deployable in prenatal care. Prior studies have shown that AI models trained on retinal images can accurately predict demographic and clinical parameters such as age, sex, and hemoglobin levels, as well as future risk of systemic diseases including diabetes, stroke, and cardiovascular disease^41–44^.

Initial efforts to apply AI to HDP prediction via retinal imaging have been promising but limited. The PROMPT model (Preeclampsia Risk factor + Ophthalmic data + Mean arterial pressure Prediction Test)^45^ relied heavily on clinical variables (e.g., MAP) and achieved modest image-only performance (AUC ∼0.6). Another study combining 45-degree retinal fundus images with clinical data reported improved accuracy, but used a convolutional neural network architecture that lacks interpretability and biological insight^46^. Notably, both studies were conducted in homogenous East Asian populations, further limiting their generalizability and translational impact.

### Visionary AI: A Mechanistically Grounded, Interpretable Framework for Risk Screening

Here we introduce *Visionary AI*, an end-to-end computational framework that takes as input ultra-widefield retinal images and predicts an individual’s risk of hypertensive disorders of pregnancy. Unlike prior approaches that treat retinal images as opaque inputs to generic deep learning models, Visionary AI represents a fundamentally different approach: it models the retinal vasculature as a quantifiable, interpretable biological system – one that mirrors systemic endothelial and angiogenic processes occurring during placental development.

We show across a cohort of 1,267 pregnant participants that this framework enables accurate and interpretable prediction of preeclampsia and gestational hypertension months before clinical diagnosis. Our results demonstrate that the maternal retina can serve as a proxy biosensor for systemic vascular health in pregnancy – opening a new frontier in non-invasive, biologically grounded, and globally scalable obstetric risk stratification.

## Results

### Cohort Characteristics and Study Design

A total of 1,267 pregnant individuals that attended their prenatal visits between the years of 2021 and 2025 at the New York-Presbyterian Hospital (Columbia University Medical Center) were recruited for this study. Within this cohort, 55 (4.3%) participants were diagnosed with preeclampsia, which was defined and diagnosed using the 2013 ACOG Guidelines^8^. Participants with preeclampsia were further stratified based on disease severity, with 38 (3.0%) classified as “with severe features” (SF), and 17 (1.3%) classified as “without severe features” (NSF). Additionally, 20 (1.6%) were classified as EOPE, while the remaining 35 (2.8%) were classified as LOPE. All cases were independently reviewed twice (maternal-fetal medicine specialists WB, and EA) both of whom were masked to the retinal imaging results. One participant was excluded from the retinal analysis due to poor image quality, reducing the preeclampsia, NSF, and LOPE case counts by one.

**Table S1** summarizes the demographic and clinical characteristics of the study population. Clinical data collected included maternal age, race/ethnicity, tobacco use, prior pregnancy history, pre-existing conditions, and medication use. We also recorded pregnancy-related complications (e.g., gestational hypertension, gestational diabetes), and birth outcomes, including small for gestational age (SGA, <10th percentile), severe SGA (<3rd percentile), stillbirth, birth weight, and Apgar scores. Notably, 85% of participants self-identified as Hispanic or Latinx. There were no significant differences between participants who developed preeclampsia and those who did not in terms of maternal age, race/ethnicity, obesity status, or tobacco use. However, individuals who developed preeclampsia were significantly more likely to have a history of preeclampsia in prior pregnancies (16.4% vs. 3.8%; p = 4.1e-05, Chi-square test) and to have pre-existing hypertension (18.2% vs. 3.5%; p = 5.5e-07, Chi-square test).

Additionally, participants who developed gestational hypertension had significantly higher rates of obesity compared to normotensive population-wide controls (65% vs. 33%; p = 6.5e-06, Chi-square test).

Retinal images were acquired using the Optos SLO Primary fundus camera during the first (6– 13 gestational weeks, GW), second (14–27 GW), and third (28–40 GW or until delivery) trimesters. **Figure 1** provides an overview of the study design and the Visionary AI computational framework, including the sample collection timeline, central hypothesis, and model architecture.

**Figure 1.**
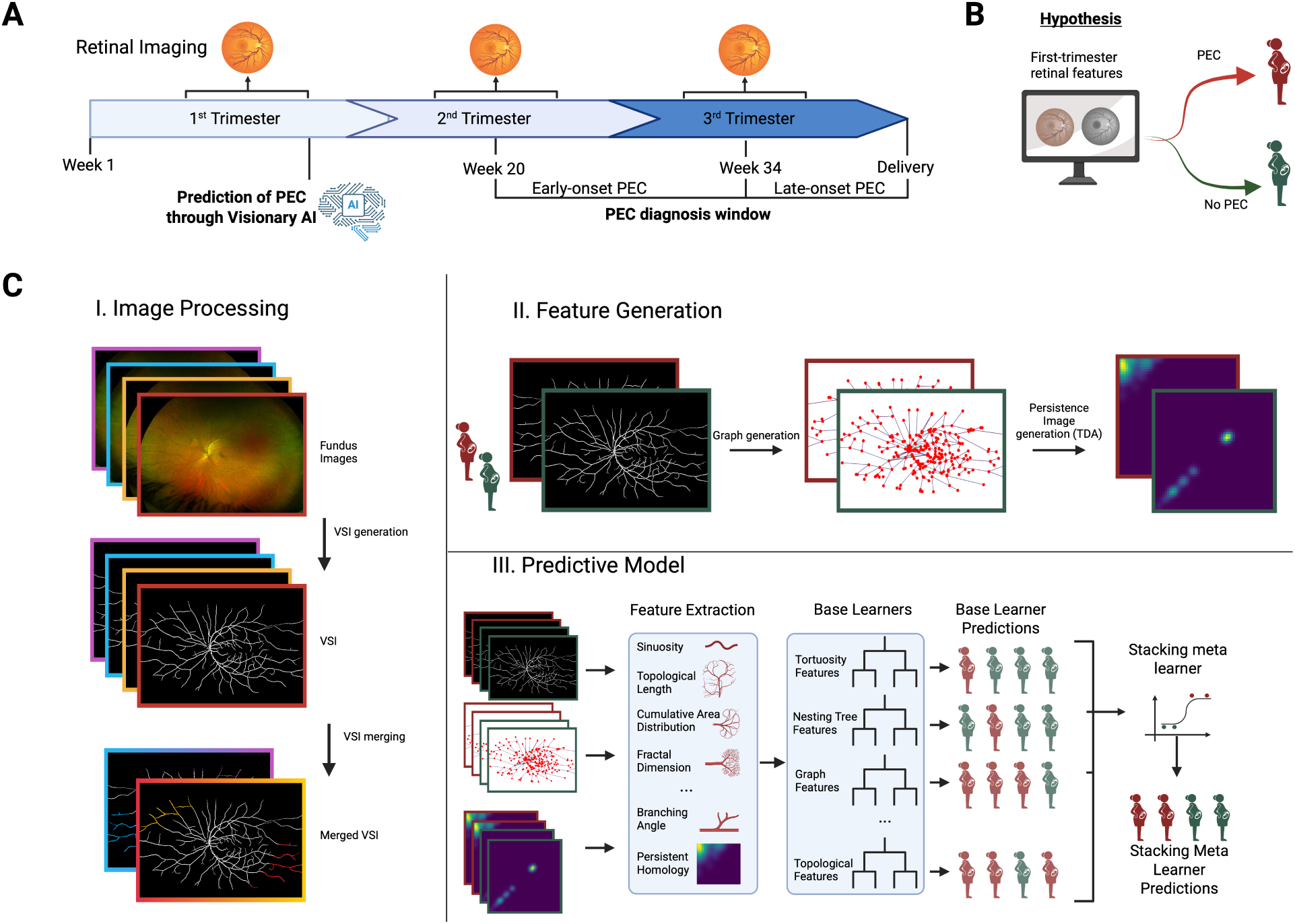
Study and Algorithm Design. **(A)** A timeline of the study procedure. Participants were approached for retinal imaging in the first, second, and third trimesters. Prediction of preeclampsia was done utilizing the first trimester images, except for participants that lacked these images, in which case second trimester images were used. Clinical preeclampsia diagnosis was given after week 20 of gestation. Preeclampsia cases are considered early-onset if the diagnosis occurred before week 34, and late onset if it occurred after week 34. **(B)** The primary hypothesis of the study is that it is possible to accurately predict the risk of developing preeclampsia using first trimester retinal images. **(C)** Our computational pipeline can be broadly divided into three modules. I. Image processing: fundus images are collected for both eyes for a given participant. VSIs are generated for each image. Images for the same eye are merged to create a more complete VSI. II. Feature generation: the VSI are used to generate informative features related to the vascular structure of the retina. III. Predictive model: we utilize an ensemble model architecture which uses base learners to make individual predictions based on groups of features. These predictions are aggregated using a stacking meta-learner to make a final prediction. *PEC (preeclampsia); VSIs (vessel segmentation images)*.

### The Visionary AI Framework

Visionary AI is an end-to-end computational framework that takes as input ultra-widefield retinal images and predicts an individual’s risk of hypertensive disorders of pregnancy, including preeclampsia (as defined in the ACOG guidelines), EOPE, LOPE and gestational hypertension. The pipeline begins with the generation of vessel segmentation images (VSIs) using a bespoke version of the retinal vessel generative adversarial network (RV-GAN)^47^. The model was initially trained on external datasets containing fundus images (45 degrees) and hand annotated VSIs to establish baseline performance^48–52^. The model was then fine-tuned using high-quality, ultrawide-field segmentation maps curated from retinal images of participants in our cohort. These maps were manually annotated and validated by a retinal specialist, ensuring both accuracy and clinical reliability (see Methods for details). For visits with multiple images per eye, VSIs were merged to create a single coherent representation per eye, accounting for variation in gaze and image acquisition. This fusion was performed in two steps: (1) feature-based homography to achieve coarse alignment of the VSIs, addressing global shifts in positioning, and (2) deformable image registration to correct local non-linear distortions in vascular geometry. The latter step was critical for aligning vessel structures that appeared warped or spatially inconsistent across frames, enabling anatomically faithful composite VSIs suitable for downstream risk prediction.

The merged VSIs were transformed into graph-based representations to enable vascular feature extraction. In this representation, nodes were assigned to vessel branch points and terminations, with edges connecting adjacent nodes to form a vessel network. From these graphs, structural features – such as node degree (e.g., number of branches per bifurcation), total number of nodes, and branching complexity – were computed. In parallel, geometric features, such as vessel tortuosity, curvature, and loop complexity, were derived either from the graph or directly from the binary VSI. To capture higher-order vascular topology, we applied topological data analysis (TDA): retinal vessel pixels were subsampled and processed to generate persistence images, which encode topological features such as connected components and loops across scales. Additionally, higher order features were extracted by analyzing the hierarchical structure of the loops in the vasculature of the retina^53^. All extracted features were subsequently grouped into semantically related feature sets. Each set was then used to train an independent classifier (base learner). These predictors serve as building blocks for the final ensemble model. We evaluated three classification algorithms as base learners: Logistic Regression (LR), Random Forest (RF), and XGBoost (XGB). Each model was trained independently on individual feature sets derived from the VSIs, with hyperparameters tuned separately for each feature set–classifier pair. These base learners were then integrated using a stacked generalization framework, wherein their out-of-fold predictions served as inputs to a XGBoost meta-learner with recursive feature elimination, which generated the final risk prediction. Both base learners and the meta-learner were trained using leave-one-out cross-validation (LOOCV), a strategy selected to optimize data utilization in the context of our prospective study design and the low incidence of preeclampsia and other HDPs. LOOCV minimizes overfitting and yields robust, unbiased performance estimates, particularly important for rare outcome prediction^54,55^.

### Prediction of Preeclampsia from Retinal Features

Visionary AI demonstrated high predictive performance using retinal images obtained in early pregnancy, achieving an achieving an area under the receiver operating characteristic curve (AUC) of 0.94, average precision (AP) of 0.92, and a positive predictive value (PPV) of 0.85 in distinguishing individuals who subsequently developed preeclampsia (n = 54) from healthy controls (n = 82; **Figure S1**). The healthy controls were defined as study participants without any background medical conditions or pregnancy complications. Specifically, the healthy control group excluded individuals with maternal comorbidities, pregnancy complications, or ocular conditions – including cardiometabolic, neurologic, vascular, hematologic, endocrine, infectious, or genetic diseases – as well as those with multifetal gestation, prior ocular surgery or trauma, medication use (e.g., antihypertensives, insulin, aspirin), or pregnancies affected by hypertensive disorders, gestational diabetes, cholestasis, hyperemesis, stillbirth, multiple gestation, or smoking (see Methods for details). This set of 82 healthy controls was selected from a larger group of 176 healthy controls in the cohort to maintain class balance for model training and to enable approximate case-control matching on key demographic and imaging variables.

To evaluate the generalizability and clinical utility of our approach, we next assessed its performance in predicting preeclampsia among a broader, population-wide group of non-cases (n = 1,083) – more reflective of real-world obstetric populations than the rigorously defined healthy control subset. In this expanded cohort, we relaxed exclusion criteria to omit only multiple pregnancies (e.g., twins/triplets) and/or pregnancies ending in stillbirth, thereby including individuals with common comorbidities such as pre-existing hypertension, diabetes, and obesity. This allowed us to test model performance in a clinically heterogeneous setting.

Using bootstrapped subsets (10 iterations, nᵢ ≥ 100), Visionary AI maintained strong predictive accuracy, achieving a mean AUC of 0.90 ± 0.06, AP of 0.76 ± 0.11, and PPV of 0.79 at a 0.5 threshold (**Figure 2AB**). These results demonstrate that model performance generalizes beyond highly selected populations, retaining predictive power even in the presence of comorbidities that can influence retinal morphology. The modest decline in performance compared to the healthy control group likely reflects increased biological and imaging heterogeneity in the population-wide cohort. Adding clinical variables to the model via an additional base learner did not significantly improve predictive performance (AUC = 0.90 ± 0.06; AP = 0.78 ± 0.10; PPV = 0.79), suggesting that the retinal imaging features alone capture most of the signal relevant to preeclampsia risk. While clinical data may offer complementary insights, they appear to contribute minimally beyond what is already captured by the retinal vasculature.

**Figure 2.**
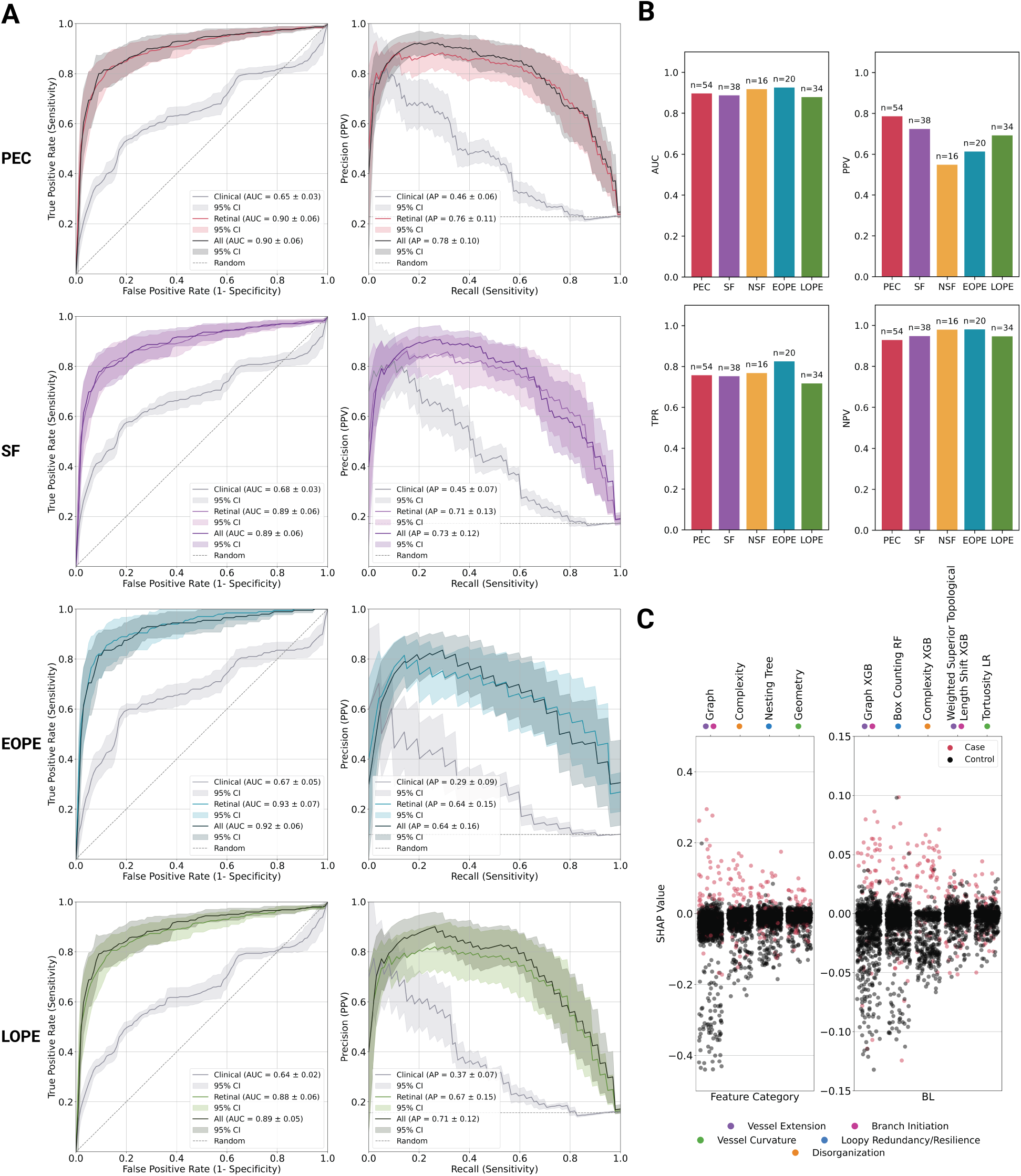
Visionary AI performance predicting preeclampsia with Population-Wide Controls. **(A)** ROC curves, PR curves, and retinal model SHAP distributions by category and key base learners for different preeclampsia subgroups. The retinal model outperforms the clinical model in all cases. The all model, which combines both retinal and clinical features performs the best. **(B)** ROC along with average Precision (PPV), NPV, and TPR values are shown for retinal model in barplots with a threshold of 0.5. FPR for all subsets of 0.06. **(C)** SHAP values for key feature categories and base learners, labeled with relevant biological processes. *PEC (preeclampsia); SF (severe features); NSF (no severe features); EOPE (early onset preeclampsia); LOPE (late onset preeclampsia)*.

### Prediction of Preeclampsia subtypes from Retinal Features

We further evaluated the performance of the model within clinically relevant subtypes of preeclampsia to ensure that overall accuracy was not driven by a single subgroup. Specifically, we assessed model performance separately for preeclampsia with severe features (SF; n = 38) and without severe features (NSF; n = 16). NSF was defined as new-onset hypertension after 20 weeks of gestation (≥140/90 mmHg on two occasions, at least 4 hours apart) accompanied by either proteinuria or mild signs of maternal organ dysfunction (e.g., mild thrombocytopenia, mildly elevated liver enzymes, or renal impairment not meeting severe thresholds)^8^. SF met the same diagnostic criteria but included at least one severe complication – such as systolic BP ≥160 mmHg, diastolic BP ≥110 mmHg, thrombocytopenia, elevated liver enzymes, renal insufficiency, pulmonary edema, or new-onset neurological symptoms^8^.

Despite the small sample sizes, Visionary AI maintained strong predictive performance for both subtypes: SF cases were predicted with an AUC of 0.89 ± 0.06, AP of 0.71 ± 0.13, and PPV of 0.72, while NSF cases achieved an AUC of 0.92 ± 0.06, AP of 0.57 ± 0.20, and PPV of 0.55. These findings confirm that Visionary AI can accurately predict both preeclampsia subtypes, while maintaining comparable predictive performance when the groups are analyzed together.

We further applied a subgroup analysis to examine the ability of Visionary AI to predict early-(EOPE: n = 20) and late-(LOPE: n = 34) onsets of preeclampsia. These subtypes are clinically defined by the timing of diagnosis – EOPE occurring at or before 34 weeks of gestation, and LOPE after 34 weeks. Emerging evidence suggests that EOPE and LOPE may arise from partially distinct biological mechanisms, with a stronger association between LOPE and maternal cardiovascular or metabolic risk factors and EOPE more associated with placental processes^9,10^. The EOPE subset was predicted with an AUC of 0.93 ± 0.07, AP of 0.64 ± 0.15, and a PPV of 0.61, while LOPE was predicted with an AUC of 0.88 ± 0.06, AP of 0.67 ± 0.15, and a PPV of 0.69, indicating high performance even when evaluating more heterogeneous clinical presentations.

As with overall preeclampsia prediction, the inclusion of clinical features provided only marginal gains in model performance across subtypes, indicating that the retinal vasculature alone captures the dominant predictive signal (SF: AUC = 0.89 ± 0.06, AP = 0.73 ± 0.12, PPV = 0.72; NSF: AUC = 0.92 ± 0.06, AP = 0.60 ± 0.19, PPV = 0.55; EOPE: AUC = 0.92 ± 0.06, AP = 0.64 ± 0.16, PPV = 0.60; LOPE: AUC = 0.89 ± 0.05, AP = 0.71 ± 0.12, PPV = 0.70).

### Benchmarking with Existing Risk Models for Preeclampsia

To evaluate the added predictive value of retinal imaging, we benchmarked the performance of Visionary AI using retinal features alone against models trained on clinical features alone, which reflect current approaches to preeclampsia risk assessment. We included a variety of clinical factors encompassing both demographics and relevant diagnostic history, including age and prior adverse pregnancy outcomes such as preeclampsia and gestational diabetes, representing a modified version of the Fetal Medicine Foundation (FMF) preeclampsia risk assessment model^56^. Several FMF variables – such as maternal height, weight, last menstrual period, and portions of maternal medical history – were unavailable in our dataset (see Methods for details).

To ensure comparability, we used the same model architecture (evaluating LR, RF, and XGB) for both the FMF clinical-only and retinal-only models. This approach isolates the impact of feature type – clinical vs. retinal – on model performance. The clinical-only model showed limited predictive ability in distinguishing cases from population-wide controls across preeclampsia subtypes (preeclampsia: AUC = 0.65 ± 0.03, AP = 0.46 ± 0.06, PPV = 0.50; SF: AUC = 0.68 ± 0.03, AP = 0.45 ± 0.07, PPV = 0.44; NSF: AUC = 0.59 ± 0.04, AP = 0.15 ± 0.06, PPV = 0.18; EOPE: AUC = 0.67 ± 0.05, AP = 0.29 ± 0.09, PPV = 0.30; LOPE: AUC = 0.64 ± 0.02, AP = 0.37 ± 0.07, PPV = 0.37). In contrast, and as reported above, the retinal-only model performed substantially better across all preeclampsia subtypes (preeclampsia: AUC = 0.90 ± 0.06, AP = 0.76 ± 0.11, PPV = 0.79; SF: AUC = 0.89 ± 0.06, AP = 0.71 ± 0.13, PPV = 0.72; NSF: AUC = 0.92 ± 0.06, AP = 0.57 ± 0.20, PPV = 0.55; EOPE: AUC = 0.93 ± 0.07, AP = 0.64 ± 0.15, PPV = 0.61; LOPE: AUC = 0.88 ± 0.06, AP = 0.67 ± 0.15, PPV = 0.69), and adding clinical features yielded minimal performance gains. This suggests that the retinal signal captured by Visionary AI already encompasses much of the predictive information provided by the clinical features.

At fixed false positive rates (FPRs) of 5%, 10%, and 15%, Visionary AI achieved substantially higher true positive rates (TPRs) and positive predictive values (PPVs) than those reported for the PROMPT model^45^, which used narrower-field RetiCam3100 fundus imaging (∼135 degrees) and incorporated maternal risk factors, mean arterial pressure (MAP), and the retinal image. Specifically, Visionary AI yielded TPRs and PPVs of 0.68 and 0.80 (5% FPR), 0.79 and 0.71 (10% FPR), and 0.84 and 0.65 (15% FPR). In comparison, PROMPT reported markedly lower TPRs and PPVs of 47.7 and 32.3, 60.5 and 23.1, and 72.1 and 19.3, respectively, at the same FPR thresholds.

It is important to note that this is not a direct, head-to-head comparison – our analysis did not use the PROMPT model or its dataset, as neither the code nor data are publicly available. These findings suggest that Visionary AI may offer markedly improved sensitivity and precision, even under strict false positive constraints, and further underscore its potential as a robust, clinically actionable tool for early detection of hypertensive disorders of pregnancy.

### Topological and Geometric Features Reflect Early Vascular Stress in Pregnancy

To interpret our model predictions using the vascular alterations preceding preeclampsia, we analyzed the vascular features extracted by the VSI pipeline. We employed both model-independent univariate analyses and model-derived Shapley Additive Explanation (SHAP) values^57^ (**Figure 2C**) to assess feature importance. The features that most consistently differentiated preeclampsia and its subtypes from both healthy and population-wide controls clustered into four distinct categories: (i) graph-topology (e.g., counts of vessel tips, branches, topological length), (ii) geometry (e.g., tortuosity, vessel caliber/thickness), (iii) complexity/organization captured by topological data analysis and box counting fractal analysis (e.g., outward radial filtrations), and (iv) nesting-tree metrics that summarize the hierarchical organization of loops and bifurcations (e.g., loops, asymmetry). **Figure 3** provides examples of vascular features that are differentially distributed across these broad categories, illustrates them graphically, and links each feature category to underlying biological processes.

**Figure 3.**
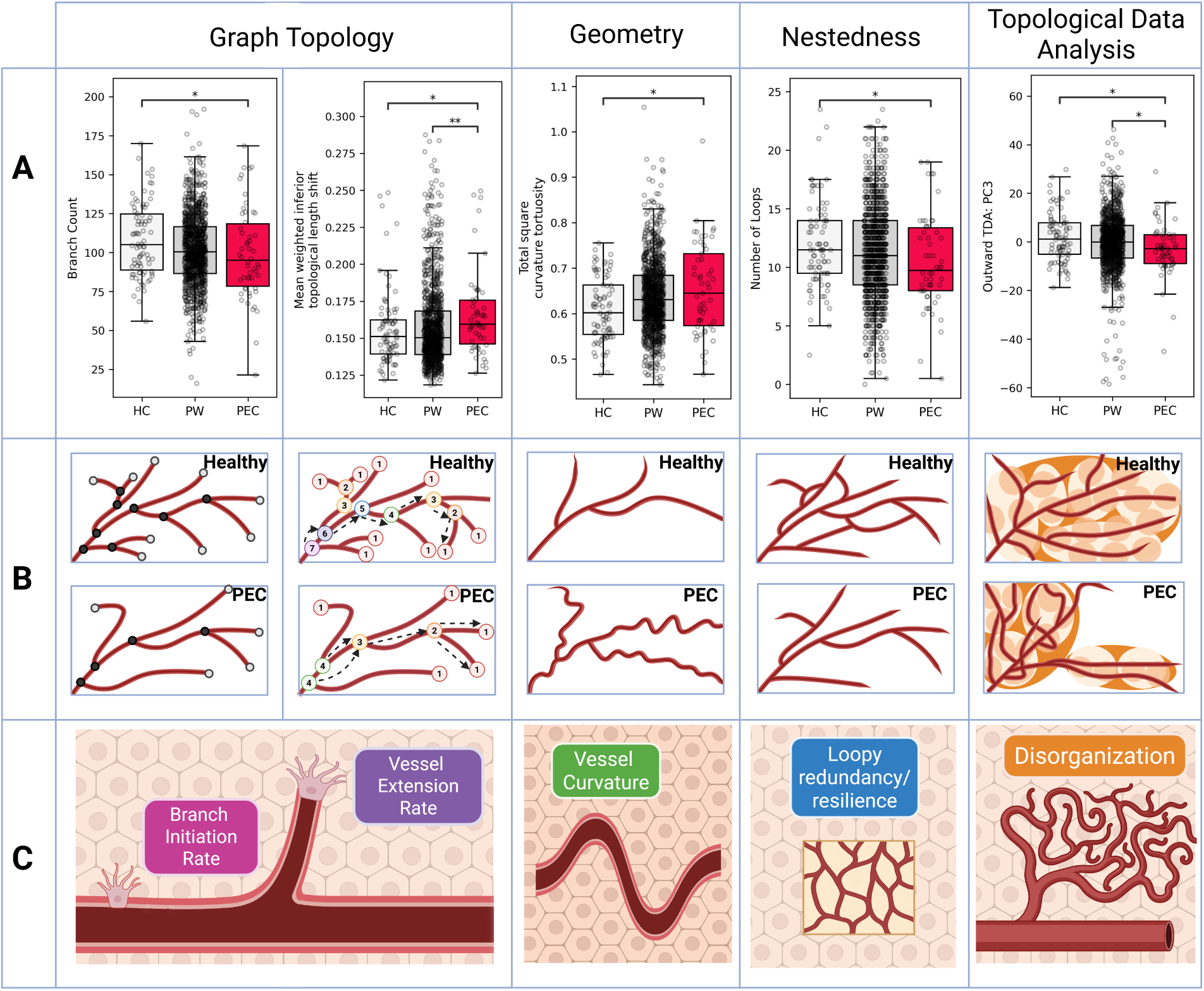
Preeclampsia-associated features reflect clinically relevant biological vessel properties. Each column reflects a feature class used to construct base learners for the ensemble model. For each class, **(A)** boxplots depicting the distributional differences between preeclampsia cases, healthy and population-wide controls across a few notable retinal features/categories, **(B)** a cartoon highlighting the differential signals captured in feature calculations within each category, and **(C)** the main relevant angiogenesis-related processes are depicted. Branch count reflects the number of branch nodes (marked with black circles in B). Topological length reflects the number of nodes between the root and the tip averaged over all branches. Tortuosity reflects the ratio between the linear distance and total vessel distance. The number of loops is a count of the loops in the vascular network. The outward radial TDA filtration captures the emergence and disappearance of connected components and loops as the vascular network is reconstructed outward from the optic disc. *PEC (preeclampsia); HC (healthy controls); PW (population-wide controls). Comparisons tested using Mann-Whitney U test. *p<0.05; **p<0.01; ***p<0.001*.

**Figure 4.**
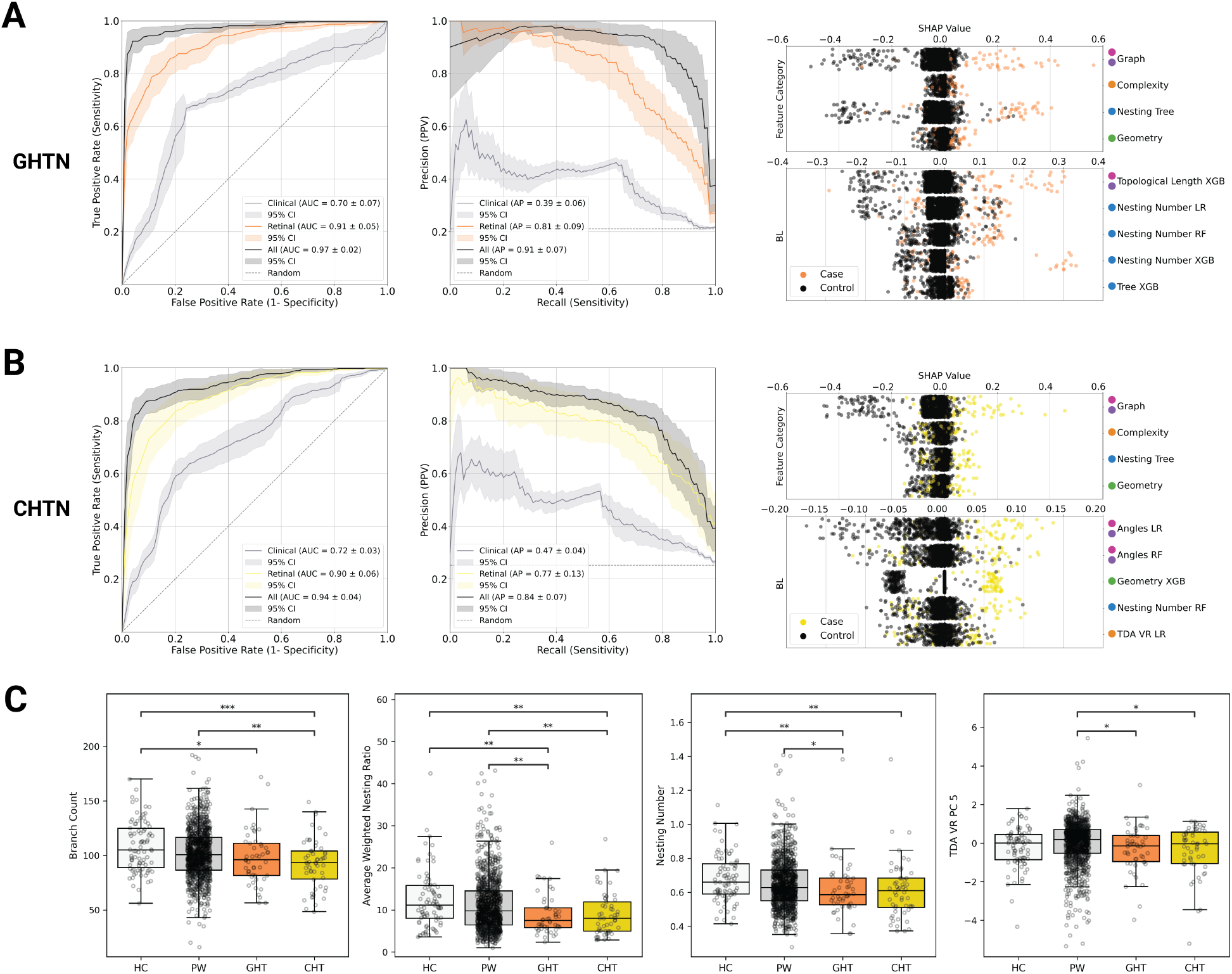
**(A-B)** ROC, PR curves, and SHAP values for prediction of gestational hypertension and chronic hypertension. The retinal model outperforms the clinical model in all cases. The combined model, which uses both retinal and clinical features performs the best for all conditions. **(C)** Boxplots depicting the distributional differences between gestational hypertension, chronic hypertension, healthy and population-wide controls across a few notable retinal features/categories. *HC (healthy controls); GHTN (gestational hypertension); PW (population-wide controls); CHTN(chronic hypertension). Comparisons tested using Mann-Whitney U test. *p<0.05; **p<0.01; ***p<0.001*.

### Univariate Analyses Reveal Early Microvascular Remodeling in Preeclampsia

Graph-topology features capture the vasculature’s connectivity (e.g., counts of vessel tips and branches, and inter-branch distances) from its graph representation. In our cohort, the number of branches was lower in preeclampsia than in healthy controls (p = 0.04, Mann–Whitney U, Figure 3A); potentially suggesting a reduced branch-initiation rate in preeclampsia. Inter-branch distances can be influenced both by branching and by tip extension (greater extension without new bifurcations tends to increase distances, whereas new branches tend to reduce them). To capture these processes jointly, we examined topological length, defined for each node as the maximum number of edges to any terminal (tip), which reflects hierarchical depth. In the inferior retina, preeclampsia cases showed a greater mean distance from both healthy and population-wide controls in the distribution of weighted topological length (p = 0.02 and 0.01 respectively, Mann–Whitney U; weighted by vessel width). Taken together, these observations suggest that preeclampsia may involve alterations in both branch initiation and vessel extension, with topological length providing a scale-free summary of their combined effects^58,59^.

Geometric features describe the shape, curvature, and caliber of retinal vessels, reflecting how efficiently blood is distributed across the vascular network. Among these, the most informative are tortuosity and vessel thickness. Tortuosity metrics quantify the extent of vessel curvature, winding, and twisting, capturing deviations from a straight path that can signal vascular stress or remodeling. We found that squared curvature tortuosity was significantly greater in samples from participants with preeclampsia and preeclampsia with SF compared to healthy controls (p = 0.01 for both, Mann–Whitney U test). However, this signal was attenuated when compared to the broader population-wide control group due to greater variability within that group. Increased tortuosity reflects enhanced vessel curvature and is often associated with altered vessel maturity, oxidative stress, and hemodynamic stress^45,60–63^. Vessel thickness, representing flow capacity, was computed as the average vessel width (see Methods for details). We observed a significant change in the distributions of vessel thickness in the superior retina of preeclampsia for SF cases compared with healthy controls (p = 0.04, Mann–Whitney U test).

To capture the network-level organization of the retinal vasculature, we quantified a set of complexity features summarizing global geometric and topological structure – including box-counting fractal analysis, TDA, and nesting tree metrics. These complementary approaches describe how vessels are spatially arranged, connected, and hierarchically branched across scales, providing a quantitative window into vascular integrity and remodeling. Box-counting complexity measures the fractal organization of the vascular tree and reflects how efficiently vessels occupy space. We observed a significant difference in the box counting distributional pattern in the preeclampsia group compared to both healthy and population-wide controls (p = 0.002 and p = 7.04e-06 respectively, Mann–Whitney U), suggesting a simpler, less space-filling vessel network consistent with microvascular loss or pruning. Notably, previous studies have also reported lower retinal fractal dimensions in women diagnosed with preeclampsia, so it is possible that we are capturing an early deviation^64^.

The outward radial TDA filtration captures the emergence and disappearance of connected components and loops as the vascular network is reconstructed outward from the optic disc. At small radii, only central trunk vessels are present; as the radius expands, peripheral branches and loops sequentially appear. This analysis summarizes central-to-peripheral vascular organization, and it revealed highly significant alterations in preeclampsia compared with both healthy and population-wide controls (p = 0.01 and p = 0.02 respectively, Mann–Whitney U). Biologically, both altered box-counting distributions and the abnormal TDA profile point to greater vascular disorganization in women with preeclampsia compared to healthy controls. Effective vascularization requires that vessel tips initiate at correct sites and subsequently migrate toward areas of greatest hypoxia to ensure uniform tissue perfusion^65,66^. This requires spatial gradients of pro and anti-angiogenic signals which, if not properly coordinated, could result in the disorganization seen in the preeclampsia patients. Coordinated pro- and anti-angiogenic signals are required to ensure that tips form and elongate in the regions they are most needed to ensure even coverage and prevent gaps or excessively overlapping vessels which alter the higher-order structure of the vessel architecture^67–69,45,70^. Disruption of these signals – as has been implicated in preeclampsia pathogenesis – could underlie the pronounced retinal vascular disorganization we quantified in early pregnancy.

The nesting tree framework further describes the hierarchical loop structure of the vasculature (following Katifori et al.^53^). Two key features – loop count and asymmetry – were significantly altered in preeclampsia eyes. We observed a reduction in the number of loops compared to healthy controls (p = 0.03 Mann–Whitney U) alongside a shift in asymmetry distribution in preeclampsia compared with population-wide controls (p = 0.006, Mann–Whitney U), potentially indicating a more tree-like and less redundant vessel network. Biologically, asymmetry quantifies how balanced the branching architecture is: lower asymmetry (a highly connected, “multiplicative” network with many loops) supports alternative flow routes and greater resilience to vessel occlusion, whereas higher asymmetry (an “additive,” tree-like structure) indicates vulnerability – where the loss of a main branch can disproportionately impair perfusion. The increase in asymmetry and loss of loops in preeclampsia retinas aligns with microvascular rarefaction and pruning observed in systemic hypertension and placental ischemia^71–74^, reflecting a shift toward a less adaptive vascular topology. For example, mid-pregnancy evaluations of systemic microcirculation have shown that reduced capillary density (structural rarefaction) independently predicts later preeclampsia, underscoring a pathologic loss of microvascular channels^75^. Our retinal findings mirror this phenomenon in the eye, with a sparser vascular network and the removal of looped collateral paths.

Of note, at the univariate level, EOPE cases had a different complexity and nesting tree profile that made it distinct from controls and from LOPE. For example, box counting distribution was significantly altered compared to both healthy and population-wide controls (p = 1e-4 and p = 2.8e-06 respectively, Mann–Whitney U, **Figure S4**), as well as compared to LOPE (p = 0.006, Mann–Whitney U). EOPE cases also had a significant reduction in the number of loops compared with both healthy and population-wide controls (p = 0.004 and p = 0.016 respectively, Mann–Whitney U). EOPE had a distinct ratio of weighted topological length in the superior retina to inferior retina compared to both healthy and population-wide controls (p = 0.02 and p = 0.01 respectively, Mann–Whitney U), as well as compared to LOPE (p = 0.02, Mann–Whitney U). This topological length signal is also present in the form of an altered distribution in the weighted topological length in the inferior retina compared to both healthy and population-wide controls (p = 0.03 and p = 0.02 respectively, Mann–Whitney U). These findings support the hypothesis – commonly suggested in the literature^9^ – that EOPE and LOPE may involve partially distinct biological mechanisms. While both conditions share clinical features, our results align with prior work suggesting that EOPE and LOPE may reflect divergent pathophysiological trajectories. Nonetheless, we can accurately predict both conditions using the Visionary AI approach.

### SHAP and Univariate Analyses Patterns Converge on Key Vascular Signatures

Graph-topology base learners exhibited the highest average absolute SHAP values when distinguishing preeclampsia from both healthy and population-wide controls, followed by complexity features for population-wide controls and nesting tree features for healthy controls (**Figures 2C & S2**). For population-wide controls, the most informative feature sets included graph topology, box counting, and vessel branching angles. These findings are consistent with univariate analyses showing that vessel extension, branch initiation, and vascular sparsity differ significantly in preeclampsia.

Other important feature sets included geometric complexity, superior topological length shifts, and tortuosity. In comparisons with healthy controls, nesting tree asymmetry (the ratio of descendants on the left and right subtrees) emerged as a particularly informative signal, highlighting the potential of asymmetric vascular architecture to distinguish preeclampsia from healthy states (**Figure S3**).

Taken together, the consistent importance of graph-based, angle-based, and box counting features across models and control groups underscores their robust predictive value. These vascular signatures also align with known retinal changes observed later in pregnancy in individuals with preeclampsia – including reduced vessel caliber and lower perfusion density – features likely reflecting systemic vasoconstriction and endothelial dysfunction that restrict microvascular blood flow. The ability of Visionary AI to detect these signatures early in pregnancy underscores both its biological validity and its potential as a powerful screening tool.

### Visionary AI Generalizability Across Gestational Hypertension and Pre-existing Hypertension

Finally, we evaluated the ability of the Visionary AI framework to predict additional hypertensive disorders, specifically gestational hypertension and chronic (pre-existing) hypertension diagnosed prior to pregnancy. Although both gestational hypertension and preeclampsia are thought to originate in placental dysfunction^76^, we hypothesized that they give rise to distinct retinal vascular signatures detectable in first-trimester images. Specifically, we expect that preeclampsia and gestational hypertension each produce unique patterns of microvascular remodeling, with preeclampsia potentially exhibiting more spatially extensive alterations. These clinical phenotypes also differ in their comorbidities: participants who developed gestational hypertension had significantly higher rates of obesity (65% vs. 33%; p = 6.5e-06, Chi-square test), while those who developed preeclampsia had higher rates of chronic hypertension (18.2% vs. 3.5%; p = 5.5e-07, Chi-square test).

To this end, we used a similar model architecture to assess our approach’s generalizability in distinguishing gestational hypertension (n = 49) and chronic hypertension (n = 61, excluding subjects with superimposed preeclampsia). These new models were able to predict GHTN with an AUC of 0.91 ± 0.05, AP of 0.81 ± 0.09, and PPV of 0.81. CHTN with an AUC of 0.90 ± 0.06, AP of 0.77 ± 0.13, and PPV of 0.77.

At the univariate level, we observed a consistent reduction in vascular branch counts across all hypertensive disorders evaluated – chronic hypertension, gestational hypertension, and preeclampsia – relative to both healthy controls (p = 7e-4,0.02,0.04 respectively, Mann–Whitney U) and population-wide controls (p = 0.004, Mann–Whitney U, chronic hypertension). Despite this shared pattern, gestational hypertension and preeclampsia exhibited distinct topological profiles. Specifically, in both gestational and chronic hypertension, the mean weighted nesting ratio and nesting number were significantly reduced compared to healthy controls (p = 0.001, 0.001 and p = 0.002, 0.005, respectively, Mann–Whitney U) and to population-wide controls (p = 0.009, 0.008 and p = 0.05, 0.06, respectively, Mann–Whitney U), indicating a simplification of hierarchical vascular organization. Furthermore, topological differences in Vietoris–Rips (VR) filtrations were also significant in both gestational and chronic hypertension relative to healthy controls (p = 0.08 and p = 0.005, Mann–Whitney U) and population-wide controls (p = 0.02 and p = 0.04, Mann–Whitney U), with a larger effect size observed for chronic hypertension. This suggests localized simplification of microvascular architecture and diminished hierarchical complexity. These findings are consistent with subtle, spatially constrained changes in vessel branching or spacing that may reflect impaired autoregulatory capacity or early microvascular rarefaction.

The SHAP values for the GHTN and CHTN models align with the univariate results, with three of the top five GHTN base learners and one of the top five CHTN base learners using the nesting number feature set (**Figure S5 & S6**). The CHTN model differs from GHTN, similar to the univariate results, with the Angles LR & RF as its top two base learners.

In contrast, preeclampsia was characterized by a broader pattern of global vascular remodeling. Specifically, we found significant differences in TDA outward filtrations, consistent with disruption of global vascular morphology and boundary organization. These changes were accompanied by a reduced number of vascular loops, alterations in asymmetry and box-counting metrics, and elevated tortuosity – collectively suggesting a loss of structural regularity and potential disruption of bilateral and scale-invariant vascular organization.

Overall, our model demonstrates strong predictive performance for both conditions, supporting the notion that retinal imaging can detect early, subclinical vascular changes in high-risk pregnancies. The differences in retinal topology may reflect the divergent timing, severity, or underlying mechanisms of these hypertensive disorders of pregnancy.

## Discussion

### Early-Pregnancy Retinal Imaging Enables Pre-Symptomatic Prediction of Hypertension Disorders of Pregnancy

Visionary AI detects retinal vascular signatures of hypertensive disorders of pregnancy from ultra-widefield imaging, enabling prediction well before clinical symptoms emerge. By detecting subtle signs of placental maladaptation and systemic endothelial stress, Visionary AI offers a biologically grounded, interpretable, and scalable solution for early risk stratification across diverse obstetric populations.

Retinal images were acquired using a 200degree Optos ultra-widefield system, providing an ∼80% view of the retina and enabling detection of early microvascular perturbations that may reflect placental dysfunction or systemic vascular stress. Within a prospective U.S. cohort of 1,267 pregnancies, Visionary AI achieved high predictive performance across preeclampsia subtypes – AUC = 0.90 for preeclampsia, 0.93 for early onset preeclampsia, and 0.89 for preeclampsia with severe features – substantially exceeding the performance of a first trimester clinical only model (AUC = 0.65). The model also generalized to predict gestational hypertension (AUC = 0.91) and chronic hypertension (AUC = 0.90), underscoring its broad clinical relevance.

### Interpretable Vascular Signatures Reflect Early Systemic Pathophysiology

A key advance of Visionary AI is its interpretable framework. Rather than relying solely on deep learning, the algorithm constructs a graph-based representation of the retinal vasculature and extracts topological, geometric, and complexity-based features from vessel segmentation images. These signatures consistently distinguished cases from both healthy and population wide controls, with graph based, angle based, and box counting feature sets emerging as the strongest contributors – patterns that align with the univariate analyses demonstrating differences in vessel extension, branch initiation, and vascular sparsity.

These retinal signatures mirror hallmark pathology observed later in pregnancy across retinal imaging studies^40,75^, including reduced vessel density, lower perfusion, diminished branching complexity, and increased vascular asymmetry. Such findings are thought to reflect a combination of anti-angiogenic signaling, microvascular rarefaction, and altered vascular tone. Notably, our model detected these signatures months before symptom onset – reinforcing their biological grounding and supporting the concept of early systemic vascular remodeling in the pathogenesis of HDPs.

### “Simplified” Vasculature as a Systemic Hallmark of Preeclampsia

Visionary AI captures early signs of vascular simplification, a hallmark of preeclampsia, driven by two complementary pathological processes: capillary network rarefaction^40,75^ (loss of small vessels, reduced density, and impaired branching complexity) and arteriolar wall thickening (hypertrophic remodeling that narrows luminal diameter and increases resistance)^77^. These processes jointly contribute to reduced network redundancy, lower perfusion, and heightened susceptibility to ischemic injury^12,14,79^. Our topological metrics, including changes in loop structure, asymmetry, and branching organization, reflect these early deviations from physiologic vascular remodeling. Superior–inferior retinal differences in feature importance further align with known variations in regional blood flow^78^. That Visionary AI identifies these shifts early in gestation underscores the system-wide vascular impairment characteristic of HDPs and provides mechanistic insight into divergent early- and late-onset disease pathways.

### Comparison to Prior AI Approaches

While recent work has explored AI-based preeclampsia prediction using retinal imaging – most notably the PROMPT model^45^ – these models have shown modest performance (AUC ∼0.6 using images alone), rely heavily on clinical variables (e.g., MAP), and were developed in homogeneous East Asian cohorts. Visionary AI differs fundamentally in design and performance: it requires no clinical inputs, is mechanistically interpretable, and achieves substantially higher predictive accuracy in a racially and clinically diverse population.

At matched false positive rates (5%, 10%, 15%), Visionary AI consistently achieved higher true positive rates and positive predictive values than PROMPT. This is not a head to head comparison – PROMPT code and data are not publicly available – but the magnitude of performance difference underscores the value of biologically informed vascular modeling combined with ultra-widefield imaging.

### Clinical Translation and Global Scalability

Visionary AI offers a fast, noninvasive, and cost effective screening strategy suitable for both high resource and low resource settings. Retinal fundus imaging requires no laboratory infrastructure, takes less than two minutes to perform, and can be operated by nonspecialists. As our computational pipeline is automated and does not require manual image grading, it eliminates subjectivity and reduces implementation barriers.

This approach is particularly promising for low and middle income countries (LMICs), where cold chain dependent biomarker assays and Doppler ultrasound are difficult to deploy at scale. Retinal imaging – already widely used for diabetic retinopathy screening globally – could be integrated into routine first trimester antenatal care to support early aspirin initiation, blood pressure monitoring, and timely referral pathways.

### Limitations and Future Directions

Several limitations warrant consideration. Although our prospective cohort was large for imaging based pregnancy research, the absolute number of early onset and severe preeclampsia cases was modest due to low baseline prevalence. The single-site design limits demographic breadth and calls for validation across diverse geographic and clinical settings. A higher-than-expected proportion of severe cases likely reflects the underlying clinical population, and rigorous masked review was performed to ensure accurate phenotyping.

Future and ongoing work includes multi-site international validation, integration with circulating biomarkers and placental pathology for deeper mechanistic insight, and longitudinal imaging to define dynamic vascular remodeling across gestation. These efforts are critical for realizing the global potential of biologically interpretable AI to transform early pregnancy care and reduce maternal morbidity worldwide.

## Methods

This prospective study was approved by the Institutional Review Boards of Columbia University and New York University and adhered to the tenets of the Declaration of Helsinki. Informed consent was obtained from every participant prior to enrollment. The study aimed to capture early retinal vascular signals in pregnancy among individuals receiving prenatal care at Columbia University, with the goal of distinguishing those who would go on to develop HDP.

### Cohort Description and Data Acquisition

Pregnant individuals receiving routine prenatal care at the New York-Presbyterian Hospital at Columbia University between 2021 and 2025 were enrolled in this study. Eligible participants were ≥ 18 years of age and able to provide informed consent. Participants could be enrolled to the study by the second trimester of pregnancy.

Maternal demographic and clinical data – including age, race/ethnicity, medical history, and pregnancy-related diagnoses such as gestational diabetes mellitus (GDM) and hypertensive disorders of pregnancy (HDPs), as well as laboratory values like hematocrit and hemoglobin A1c – were extracted from the electronic health record. Delivery outcomes, including small for gestational age (SGA), stillbirth (SB), birth weight, and Apgar scores, were also recorded. Diagnoses were made by participants’ obstetric clinicians during routine prenatal care and cases were subsequently reviewed twice in masked fashion (WB and EA).

Retinal images were obtained for each eye by trained research technicians during the first (6–13 weeks), second (14–27 weeks), and third (28–40 weeks or until delivery) trimesters using the Optos UWF Primary scanning laser ophthalmoscope, which captures ultra-widefield images (200-degree field of view, covering over 80% of the retina). History of ocular conditions and surgery was also captured and used for inclusion and exclusion criteria. This study focused on prediction of hypertensive disorders early in pregnancy. First trimester images were used when they were available, otherwise second trimester images were used (32% of preeclampsia, 24% of gestational hypertension, 38% of cardiac hypertension, 43% of healthy controls, and 40% of population-wide control participants). However, even second trimester images were obtained before week 20 of gestation.

### Image Preprocessing and AI-Generation of Vessel Segmentation Images

We analyzed a total of 3,909 retinal images. Images were rescaled to a uniform zoom level and resized to a standard resolution of 912 × 912 pixels. To minimize interference with downstream processing, each image was cropped using a standardized elliptical mask to remove non-retinal artifacts, such as eyelashes or peripheral obstructions outside the field of view. This process is done by applying a uniform and programmatic cropping method.

Vessel segmentation images (VSI) were generated from these cropped fundus images using a dedicated AI model designed for retinal vessel segmentation, inspired by generative adversarial networks (RV-GAN)^47^. The architecture includes a U-Net generator that learns both the overall vessel structure and fine details. The model was trained on standard 45 degree color fundus photographs paired with expert-annotated VSIs from publicly available datasets^48–52^. The model was then tuned on a curated set of ultra-widefield (200 degrees) retinal images from our cohort (n = 20), each with corresponding hand-annotated VSIs. These initial wide-field vessel maps were independently reviewed by a retinal specialist (SB) to ensure accuracy.

The generated VSIs were binarized using a threshold of 0.3 and processed to exclude any small, disconnected components. As the model produces non-binary images, where each pixel is assigned a probability of being a vessel, 0.3 was chosen instead of the more standard 0.5 in order to detect more detail. Next, VSIs from different images of the same eye were merged together in order to produce a more complete image of the retinal vasculature. Feature matching for merging was performed using ORB (Oriented FAST and Rotated BRIEF) to detect rotation- and scale-invariant keypoints and compute binary descriptors for efficient matching; a homography was then estimated with RANSAC (Random Sample Consensus) to scale and align the images without local deformation. Then, a gradient descent image registration algorithm was used to find the transformation that resulted in the highest amount of overlap between the vessels of the two VSI being merged by locally distorting the images. In the case of more than two VSI of the same eye being merged, one VSI was chosen as the first based on order of photograph capture, and additional VSI were merged on top of it sequentially. This process was done per eye for each patient. When merged VSIs were available for a patient’s right and left eye, we extracted features from each eye independently and averaged them. If only one eye’s VSI was available, we used that eye’s data alone; monocular imaging data was used for a total of 34 patients.

### Feature Generation, Categories, and Definitions

We generated 99 base learners by training three different models (Logistic Regression; LR, Random Forest; RF and XGboost; XGB) on 33 feature sets. These base learners were then aggregated into a stacked generalization meta-learner. Base learners fall within the following broad categories: graph topology, geometry, complexity/organization, nesting tree, composite, and clinical features.

### Graph-topology base learners and features (#1-8 base learners)

Graph representations of the vessel segmentations were generated by skeletonizing the binary vessel segmentation images and tracking the vascular structure. Nodes were placed at points where vessels bifurcated or terminated, and edges represented the vessel segments connecting these nodes.

#### Feature Set #1: Graph

From each resulting graph, we extracted key structural features, including the total number of nodes (encompassing both bifurcation and terminal points), the number of branching points where vessels diverged, the number of terminal nodes corresponding to vessel endpoints, the number of disconnected components/subgraphs, the direct distance computed as the average length of an uninterrupted blood vessel segment and the mean Chebyshev distance between connected nodes along each vessel segment computed as: *D_chebyshev_* = *max* (| *x1*, *x2*|, |*y1*, *y2*|).

Chebyshev distance was chosen as the baseline because the length of a vessel cannot cover the distance between two points in a smaller number of pixels.

- Number of nodes: sum of number of bifurcation points and number of termination points
- Number of components: Total number of connected components in the vasculature
- Number of termination points
- Number of bifurcation points
- Average distance between nodes

#### Feature Set #2-3: Topological Length (superior & inferior/whole graph)

Topological length features were calculated using the graphs generated from the binary vessel segmentation images. Two versions of topological length were computed: a simple count of downstream branches until no narrower continuation was found, and a weighted version, which weighs each branch by the cross sectional width of the vessel at that point. Furthermore, the topological length was analyzed separately for the superior and inferior regions of the retina, which is relevant because dependent, or gravitational, changes may be seen in the inferior half.

- Average topological length (weighted/unweighted)
- Median topological length (weighted/unweighted)

#### Feature Sets #4-7: Weighted/Unweighted Superior/Inferior Topological Length Shift

To assess distributional differences across all vessels – not just summary statistics (e.g., mean/median) – we computed two-sample Kolmogorov–Smirnov (KS) tests, comparing each participant’s empirical distribution of topological lengths to the distribution from each healthy control. The resulting KS *D* statistics formed a feature vector (one feature per healthy control).

During leave-one-out cross-validation, the held-out control’s comparison was excluded to prevent information leakage. KS statistics for weighted and unweighted topological length were calculated for the superior and inferior vasculature.

#### Feature Set #8: Topological Length Shift

KS statistics for unweighted topological length were calculated for the whole vasculature.

### Geometry base learners and features (#9-15 base learners)

Geometric features characterize vessel shape and layout, including branching angles, vessel length, and density. Geometric features included angles, cross-sectional width of vessels, and tortuosity (a blood vessel’s abnormal twisting, bending, or winding path, deviating from its normal straight or gently curved course).

#### Feature Set #9: Geometry

- Vein density length: Sum of vessel segment lengths divided by the area of the largest enclosed loop
- Mean vein distances: Average radial distance from loop centers to their enclosing vessels
- Mean areole area: Average area enclosed by vessels (i.e., loop areas)
- Areole density: Number of loops normalized by the area of the largest loop

#### Feature Set #10: Angles

Refers to vessel branching angles at bifurcation points

- Average angle
- Median angle
- Sum angle
- Beta (bimodality coefficient)

#### Feature Set #11: Tortuosity

Several measures of tortuosity were calculated following Hart et al.^80^. Sinuosity, distance inflection count tortuosity, tortuosity density, linear regression tortuosity, square curvature and absolute curvature tortuosity. All tortuosity features are computed per vessel segment and summarized at the subject level.

- Sinuosity: Average of all segment-level sinuosity values, where each is calculated as 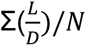 where L is the path length that the vessel follows, D is the Chebyshev distance between the two nodes at the ends of the vessel, and N is the total number of vessels. (reflects the average deviation from a straight line)
- Distance inflection count tortuosity: Sinuosity, weighted by the number of inflection points per vessel is calculated as Σ(S*I_n_)/N where S is the sinuosity of the vessel, I_n_ is the number of inflection points on the vessel, and N is the total number of vessels
- Tortuosity density: Summed relative curvature between inflection points, normalized by path length and averaged across vessels, calculated as 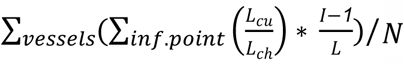 where L_cu_ is the curve length between those inflection points, L_ch_ is the chord length between those inflection points, I is the total number of inflection points on that vessel, L is the curve length of that entire vessel, and N is the total number of vessels
- Linear regression tortuosity: The average r2 value from a linear regression performed on a subset of points from each vessel is used as a measure of the linearity of the vessel, as it indicates how many points on the vessel deviate from the line of best fit
- Square curvature tortuosity: Average of squared curvature values across segments, highlighting accumulated turning severity, calculated as 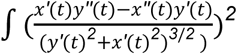, which is the square of the curvature of the vessel, integrated over every point in the vessel
- Absolute curvature tortuosity: Average of absolute curvature calculated as 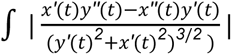, which is the absolute value of the curvature of the vessel, integrated over every point in the vessel

#### Feature Sets #12: Vessel Thickness

Vessel thickness reflects flow capacity and is calculated as the average width of each vessel. Similarly to topological length, vessel thickness was analyzed separately for the superior and inferior regions of the retina, which is relevant because dependent or gravitational changes may be seen in the inferior half.

- Average thickness (superior/inferior)
- Median thickness (superior/inferior)

#### Feature Sets #13-15: Superior/Inferior/Whole Vessel Thickness Shift

To examine distributional differences, KS statistics were computed comparing the distribution of the vessel thickness of each participant to the distributions of the healthy controls for the superior, inferior, and whole vasculature. The statistic comparing to each healthy control becomes a feature, such that there are as many features as healthy controls. Controls that are considered test cases during LOOCV are not used to calculate features that require KS statistics for the training set.

### Complexity/organization base learners and features (#16-21)

Complexity features aim to capture the complexity of the vasculature by examining how vascular structures are maintained across different scales. This is done primarily via fractal dimension and the box counting vector, as well as the different topological data analysis (TDA) filtrations.

TDA was employed to capture the geometric and structural properties of the retinal vasculature across multiple spatial scales. To achieve this, we applied four distinct persistent homology filtrations: Vietoris–Rips, inward radial, outward radial, and flooding. Each filtration produced a corresponding persistence diagram, which was then converted into a persistence image – a stable, vectorized representation of topological features across the filtration. These persistence images were subjected to principal component analysis (PCA), and the first ten principal components were retained as input features for downstream predictive modeling.

#### Feature Set #16: Box Counting

The box counting vector divides the image into a grid and counts how many squares in the grid contain vasculature. The relationship between the scale of the grid and the number of occupied squares defines the fractal dimension. The box counting vector matrix was also subjected to principal component analysis (PCA), and the first ten principal components were retained as input features for downstream predictive modeling.

#### Feature Set #17: Complexity

- Number of loops
- Fractal dimension

#### Feature Set #18: TDA Outward filtration

An outward radial filtration refers to a method of progressively building the vascular network outward from a central point, typically the optic disc. At small radii, only vessels closest to the disc are included, and as the radius increases, progressively more peripheral structures are added, allowing the capture of how vessel connectivity and loops emerge and disappear as the network expands. This filtration emphasizes the organization of retinal vessels radiating from the center toward the periphery, making it particularly relevant for identifying structural patterns that reflect normal vascular development versus pathological remodeling. Compared with the complementary inward radial filtration (which grows from periphery inward), the outward radial approach highlights central-to-peripheral organization and can reveal how disease states such as preeclampsia, diabetes, or retinopathy alter branching, coverage, or radial distribution of vessels across the retina.

#### Feature Set #19: TDA Vietoris-Rips (VR)

Points were subsampled from the vasculature binary image. Circles of increasing radius are drawn around each point. Points are connected when their circles overlap. The birth and death of topological features (e.g., loops, connected components) are tracked as the radius grows.

#### Feature Set #20: TDA Inward filtration

Starting from the most central point in the vascular graph, the radius of a large enclosing circle is gradually reduced. As points are crossed by the circle, they are connected to other points outside the circle, recreating the original graph.

#### Feature Set #21: TDA Flooding filtration

Nearby points are initially connected and then points near each complex are incorporated in it as well as the radius increases. We record at what radius length features (loops and connected components) appear, and at what radius they cease to exist. The difference from VR filtration is that the connecting radius is projected from the edges of the simplex, not only from the points.

### Nesting Tree base learners and features (#22-29)

A graph representation of the nesting tree was constructed to represent the hierarchical structure of loops and bifurcations in the vasculature, following Katifori et al.^53^. The nesting tree is constructed by representing each loop in the vasculature as a node. Loops separated by the thinnest vessels are consecutively merged, joining in the tree into a single parent node. In this way, a binary tree is constructed that merges all loops. From the resulting tree, we extracted multiple features that characterize the underlying vascular architecture. Features include counts, ratios, and asymmetry across subtrees.

#### Feature Sets #22-23: Nesting Number (full graph) & Nesting Num (nesting tree)

The nesting ratio at each node is defined as the number of descendant nodes in the left subtree divided by those in the right subtree. The resulting distribution of nesting ratios and weighted nesting ratios (weighted by the total number of descendants of this node) describes local asymmetry across the tree. These distributions were further summarized at the subject level into global metrics: the unweighted nesting number (the average nesting ratio across nodes) and the weighted nesting number (the average nesting ratio weighted by subtree size). Additionally, to examine distributional differences, KS statistics were computed comparing the asymmetry distribution of each participant to the distributions of the healthy controls. The statistic comparing to each healthy control becomes a feature, such that there are as many features as healthy controls. Controls that are considered test cases during LOOCV are not used to calculate features that require KS statistics for the training set.

- Average of the nesting ratios and sum nesting ratios where nesting ratio *r*_n_ at a node n is defined as:

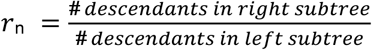 Subtree weight (*w*_n_) is *w*_n_ = *d*_n_ − 1 where *d*_n_ is the subtree degree (i.e., total number of descendant leaf nodes under *n*)
- Unweighted Nesting Number (*I_u_*) The average of all node-level nesting ratios: *I*_u_ = (1 / *N*) × Σ *r*_n_, where N is the total number of nodes
- Weighted Nesting Number (*I*_*w*_) The average nesting ratio, weighted by subtree size:

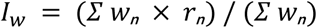

#### Feature Set #24: Tree

- Tree depth : Max branching depth of the nesting tree
- Tree leaves : Total number of leaf nodes in the nesting tree

#### Feature Set #25: Asymmetry

Tree asymmetry measures the imbalance in the number of branches between descendant nodes across all bifurcations, computed per node and summarized over subtrees of size *δ* within a window *Δ*. Asymmetry at a node (a_n_) is defined as:

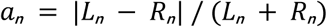

where L_n_ and R_n_ are the number of nodes in the left and right subtrees of node n, respectively, and the average asymmetry across subtrees of size *δ* is computed as:

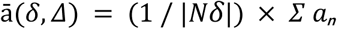

where N*δ* is the set of nodes with subtree size within *Δ*/*2* of *δ*.

- Average asymmetry, sum asymmetry, asymmetry quartiles: Aggregated branching asymmetry, based on average asymmetry per subtree of size *δ*

#### Feature Set #26: Asymmetry Shift

KS statistics for asymmetry were calculated.

#### Feature Set #27: Cumulative Size Distribution

Measures how the areas of enclosed loops in the blood vessel network are distributed. From the set of enclosed loop areas {*a*_1_, *a*_2_, …, *a*_*N*_} extracted from the nesting tree, the cumulative proportion P(a) of loops with area less than or equal to a is computed as:

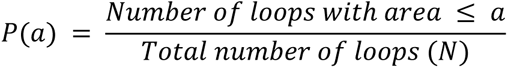

- Average cumulative size distributions, sum cumulative size distributions, cumulative size distribution quartiles : Summary statistics of cumulative proportion *P*(*a*)

#### Feature Set #28: Strahler order features

Strahler order quantifies the branching hierarchy by assigning an order of 1 to terminal branches. When two branches of the same order merge, the resulting branch is assigned an order one higher; if the merging branches have different orders, the resulting branch takes the maximum of the two. This metric reflects the structural complexity and depth of the vascular network. Bifurcation ratio captures the branching pattern by comparing the consecutive Strahler orders.

- Strahler order counts 1 to 5: Number of nodes with a given Strahler branching level, which represents how many branching points away that node is from a leaf node

#### Feature Set #29 : bifurcation ratio features

The bifurcation ratio features indicate the ratio of the number of nodes of a particular Strahler order to the number of nodes of the next Strahler order.

- Bifurcation ratio 2 to 3: Ratio of Strahler orders between levels (i.e. bifurcation level n represents Strahler-order n / Strahler order n+1

### Composite base learners and features (#30-32)

#### Feature Set #30: Ratio

Ratios that compare vascular characteristics between superior and inferior regions of the retinal image. Specifically, it includes the average and median of the superior-to-inferior ratios for topological length and for vessel thickness, calculated using both weighted and unweighted measurements.

- Average or median of superior-to-inferior ratio of topological length (both weighted and unweighted)
- Average or median of superior-to-inferior ratio of vessel thickness

#### Feature Set #31: Standard Deviation

The standard deviation of the vascular features to capture variability within each image:

- Cumulative size distribution
- Topological length (superior/inferior and weighted/unweighted)
- Vessel thickness (superior/inferior)
- Nesting ratio (weighted/unweighted)
- Angles

#### Feature Set #32: Mixed

Includes several features from different base learners described above to allow for interaction effects among predictive base learners.

- Number of branches
- Distance between branches
- Square curvature tortuosity
- Number of loops
- Fractal dimension
- Average topological lengths (superior/inferior)

### Clinical base learner and features (#33)

#### Feature Set #33: Clinical

Clinical features included maternal demographic characteristics, reproductive history, and preexisting or pregnancy-related morbidities. Specifically, we extracted data on maternal age, race and ethnicity, smoking status, and use of in vitro fertilization (IVF). We also recorded indicators of maternal risk, including advanced maternal age (AMA), prior history of hypertension, diabetes, or preeclampsia, as well as use of low-dose aspirin during pregnancy. These features comprised the clinical feature set used in downstream analyses.

### FMF benchmark model

We additionally trained a model based on the FMF algorithm^27^. This model includes features used by the FMF algorithm: maternal age, smoking status, prior history of preeclampsia or preterm labor, use of IVF, diabetes status (as well as type and medication), maternal race and ethnicity, nulliparous status, as well as preexisting hypertension. Due to lack of availability, we did not include maternal height or weight, last menstrual period and estimated delivery date, as well as limited other maternal medical history, such as vanishing twin syndrome, family history of preeclampsia, systemic lupus erythematosus, anti-phospholipid syndrome, inter-pregnancy period, gestational age at delivery and birth weight for last pregnancy. Available features were used to train a logistic regression, random forest, and XGBoost model, and the highest performing model was selected.

### Capturing distributional differences using the Kolmogorov-Smirnov (KS) Test Statistics

Some features, such as topological length or asymmetry, generate a value per blood vessel segment, resulting in a distribution for each subject. To incorporate these in our models, we first extracted summary statistics such as the median value or the quartiles. Additionally, we used Kolmogorov-Smirnov (KS) statistics to capture distributional differences: for each subject, we compared their distribution to that of a reference healthy control group. The KS statistic quantifies the maximum difference between the cumulative distribution functions (CDFs) of the two groups, with a value of 0 indicating identical distributions and higher values indicating greater divergence, effectively measuring how much a participant’s vessel-level feature distribution deviates from that of healthy controls. This method generates a feature per control. This resulting matrix undergoes a PCA decomposition, and the first ten principal components were used as the predictive features.

### Cohort Stratification

Cases were predicted against different control or non-cases groups to ascertain if the captured signal is a general disease signal (case vs healthy control), or a signature specific to preeclampsia (case vs non-case/population-wide).

The healthy control group excluded individuals with any maternal comorbidities, pregnancy complications, or ocular conditions, including cardiometabolic, neurologic, vascular, hematologic, endocrine, infectious, or genetic diseases; multifetal gestation; ocular surgery or trauma; medication use (e.g., blood pressure medications, insulin, or aspirin); and pregnancies affected by hypertensive disorders, gestational diabetes, cholestasis, HELLP, hyperemesis, stillbirth, multiple pregnancy (e.g., twins/triplets), or smoking.

For the population-wide control group, we relaxed our criteria to only exclude the following conditions relevant to hypertensive disorders of pregnancy: multiple pregnancy (e.g., twins/triplets), gestational hypertension, HELLP syndrome, and stillbirth.

### Predictive Model

A stacked Ensemble Model is used to integrate multiple feature sets representing diverse aspects of the retinal vasculature. This stacking approach improves classification accuracy by leveraging the complementary strengths of the individual models.

First, base learners were constructed by training three classifiers – Logistic Regression (LR), Random Forest (RF), and XGBoost (XGB) – on each feature set. Leave-one-out cross-validation (LOOCV) was used to optimize the hyperparameters for each base learner (LR – penalty: L1, L2; C: 0.01, 0.1, 1.0, 10; solver: saga. RF – number of estimators: 100, 200; max_depth: 1, 3, None; min_samples_leaf: 1, 2; min_samples_split: 2, 5; max_features: sqrt, 0.5, None. XGB – number of estimators: 100, 250; max depth: 1, 2, 3; learning rate: 0.01, 0.1, 0.3; subsample: 0.8, 1.0; scale positive weight: 2,8,12), with the best combination selected based on F1 score. Base learners with AUC less than or equal to 0.5 were eliminated from meta-learner consideration.

The out-of-fold predictions from the base learners were used to train an XGB meta-learner with recursive feature elimination (RFE) for base learner selection. Similar to the base learners, the meta-learner had its hyperparameters (number of estimators: 100, 250; max depth: 1, 2, 3; learning rate: 0.01, 0.1, 0.3; subsample: 0.8, 1.0; scale positive weight: 2, 8, 12; number of base learners to select with RFE: 8, 16, 32), tuned using LOOCV to prioritize F1 scores.

The healthy control ensemble model was trained on the preeclampsia cases (n = 54) and healthy controls (n = 82). This model serves to help inspect signals discriminating between cases and controls because the greatest difference is expected when comparing to a healthy cohort based on univariate analysis.

Each population-wide ensemble model was trained on the preeclampsia cases (n = 54), healthy controls (n = 82), and 10 different sets of population-wide non-healthy controls (10 times; n≥100). Performance metrics (FPR, TPR, PPV, NPV) were calculated per model and averaged. The 95% confidence intervals were determined using the standard error of the mean. This method, utilizing several population-wide sets instead of upsampling cases, was selected to specifically mitigate the chance of overfitting due to the low prevalence of preeclampsia. By performing LOOCV across 10 sets with distinct population-wide non-health controls, the confidence intervals serve as a measure of model generalizability.

For interpretability, the SHAP values are calculated for each held-out patient. Each patient’s vector of SHAP values was normalized per prediction and averaged across models for population-wide ensemble models. To calculate category SHAP, the base learner normalized and averaged SHAP values were summed per patient. Base learner selection for the meta-learners is inherently reflected in the SHAP values, with RFE excluded base learners having a SHAP value of 0.

### Univariate Analysis

The extracted features were analyzed in a univariate way to identify how individual features are affected by the disease. For each feature, the following analyses were conducted: Mann-Whitney U test, KS test, odds ratio, fold-change risk ratio, and logistic regression.

The Mann-Whitney U test is a non-parametric test used to test if for a given feature that case and control populations share the same distribution. Similarly, the KS test is also a non-parametric test used to test if case and control populations share the same distribution. It evaluates any difference in distribution, whereas the Mann-Whitney U test evaluates if there is a shift in the median.

## Data availability

Following publication in a peer-reviewed journal, we will make a curated, de-identified dataset publicly available through a secure data repository, in full compliance with applicable data sharing policies, institutional requirements, and ethical guidelines, including those governing human subjects research. Access will be provided in a manner consistent with participant confidentiality and regulatory standards. Summary-level data supporting the key findings are included in the main text and Supplementary Information.

## Acknowledgments

We thank Sabine Bousleiman, Noelle Pensec, David Wentsler, Yessenia Gutierrez, Zoilo Castillo, Jose Castillo Camacho, Casandra Almonte, Cynthia Masson, Alexa Kaminsky, Andrea P Moscoso, Ruddys Pena Recio, Emely Carmona Reyes, Desiree Torres, Stefania Maruri, Kaveri Thakoor, Ye Tian for their invaluable contributions for this work.

## Supplementary Materials

**Table 1.**
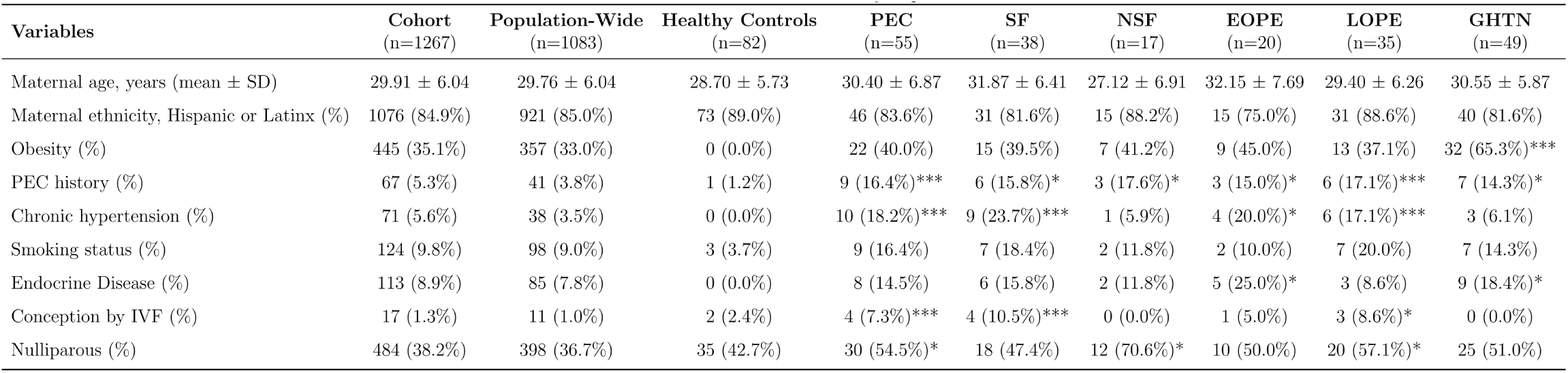
Characteristics of the study population. Clinical characteristics of study participants are presented across key population subsets and preeclampsia subtypes. The cohort-wide column includes all enrolled individuals. Population-wide controls refer to participants without major pregnancy complications in current pregnancy, excluding those with multiple gestations (e.g., twins/triplets), gestational hypertension, or stillbirth. Healthy controls represent a more stringently defined subcohort, excluding any maternal comorbidities, pregnancy complications, or ocular conditions; they comprise a subset of the 176 healthy controls identified within the full cohort. There is a subset of 80 participants who are not HDP cases but were excluded from the control group due to one or more of the following: multiple pregnancy (e.g., twins/triplets), stillbirth or a lack of usable images. *PEC (preeclampsia); SF (severe features); NSF (no severe features); EOPE (early onset preeclampsia); LOPE (late onset preeclampsia), GHTN (gestational hypertension). Comparisons tested using Mann-Whitney U test. *p<0.05; **p<0.01; ***p<0.001*.

**Supplementary Figure 1.**
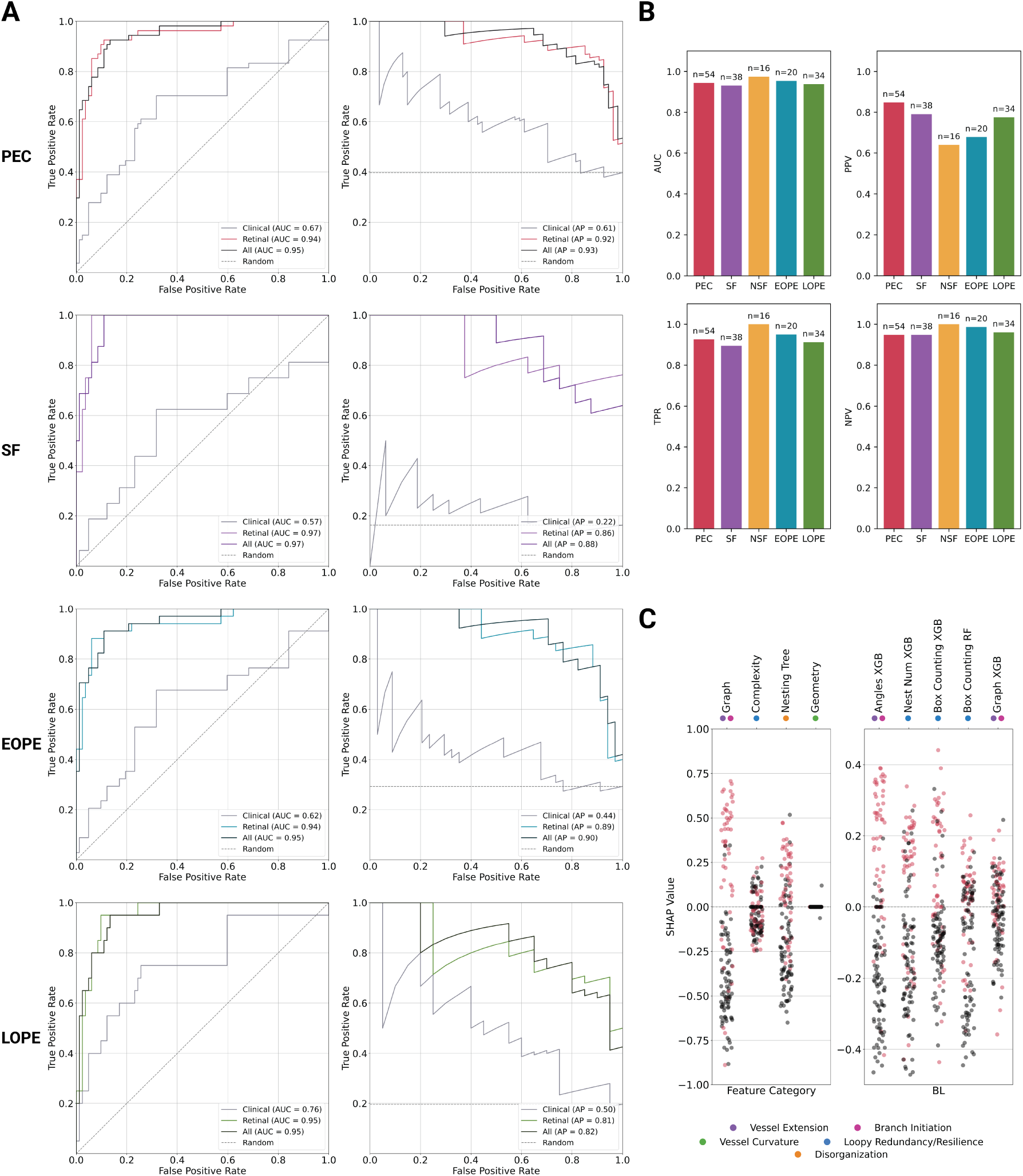
Visionary AI performance predicting Preeclampsia with Healthy Controls. **(A)** ROC curves, PR curves, and retinal model SHAP distributions by category and key base learners for different preeclampsia subgroups. The retinal model outperforms the clinical model in all cases. The all model, which combines both retinal and clinical features performs the best. The EOPE ROC and PR plots include curves for a model trained specifically for EOPE detection from retinal features. **(B)** ROC along with average Precision (PPV), NPV, and TPR values are shown for retinal model in barplots with a threshold of 0.5. FPR for all subsets of 0.11. **(C)** SHAP values for key feature categories and top five base learners based on average absolute SHAP value, labeled with relevant biological processes. *PEC (preeclampsia); SF (severe features); NSF (no severe features); EOPE (early onset preeclampsia); LOPE (late onset preeclampsia)*.

**Supplementary Figure 2.**
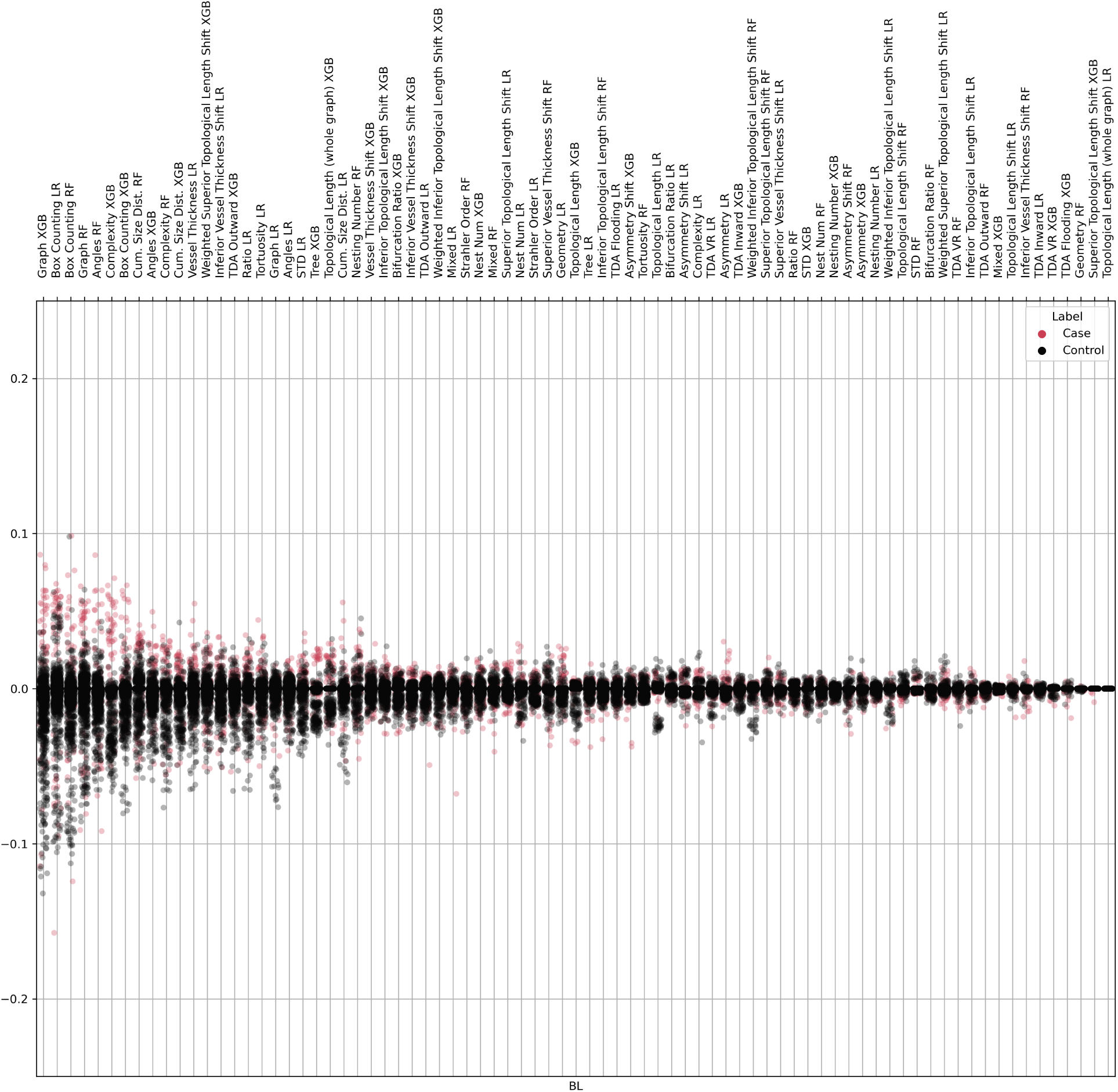
Visionary AI base learner SHAP values for predicting Preeclampsia with Population-Wide Controls. For each population-wide model, SHAP values were calculated for each patient. These SHAP vectors were normalized and averaged by patient across models. The base learners are sorted by average absolute SHAP value.

**Supplementary Figure 3.**
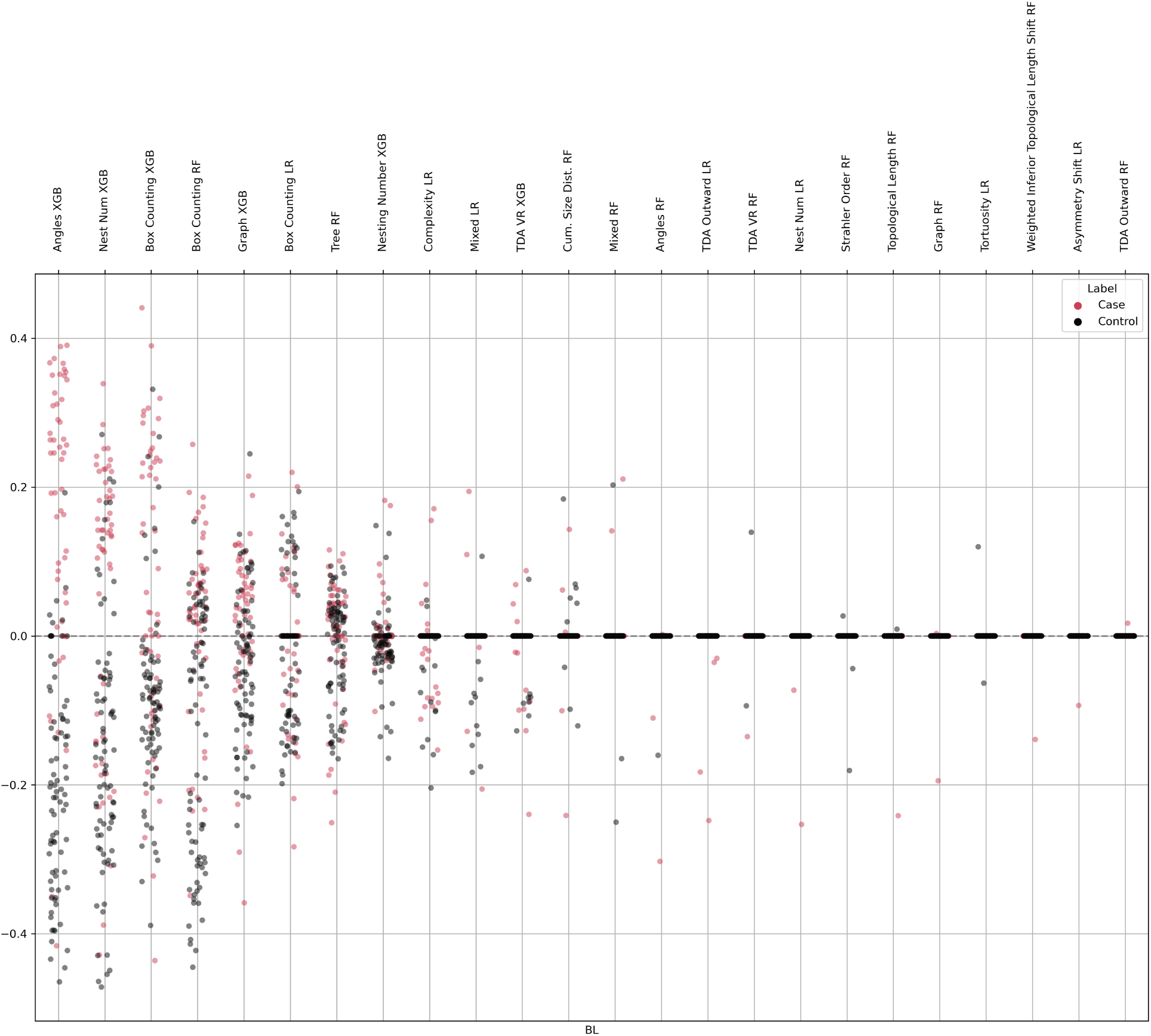
Visionary AI base learner SHAP values for predicting Preeclampsia with Healthy Controls. SHAP values were calculated for each patient. These SHAP vectors were normalized by patient. The base learners are sorted by average absolute SHAP value.

**Supplementary Figure 4.**
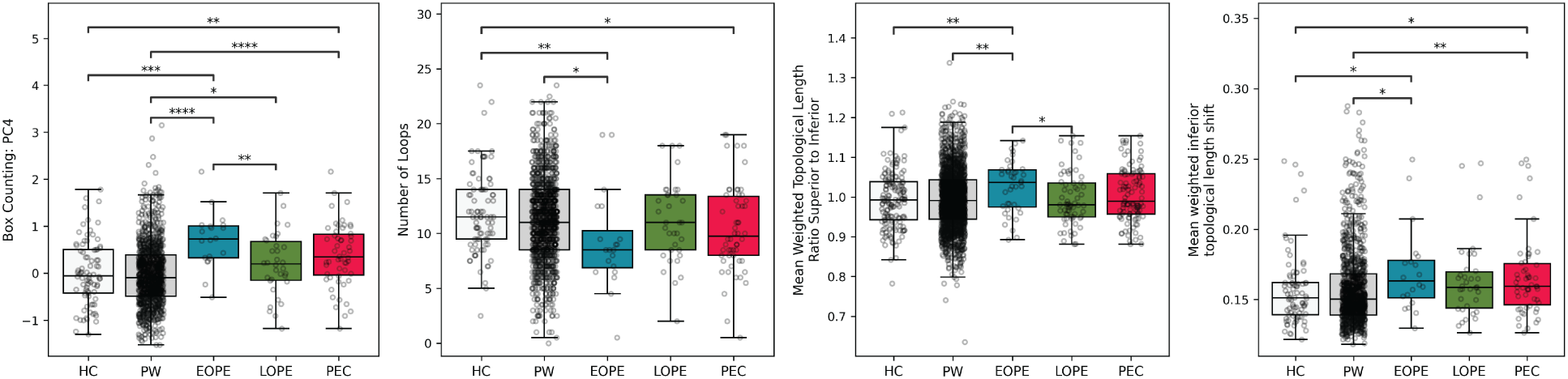
Features distinguishing early- and late-onset preeclampsia (EOPE vs. LOPE). Univariate analyses were performed to compare EOPE with both healthy and population-wide controls. These boxplots highlight features that are significantly associated with EOPE and illustrate how these patterns differ from those observed in LOPE. Together, they underscore the distinct vascular complexity and remodeling profiles of EOPE compared to other groups. *HC (healthy controls); PW (population-wide controls); EOPE (early-onset preeclampsia); LOPE (late-onset preeclampsia); PEC (preeclampsia); Comparisons tested using Mann-Whitney U test. *p<0.05; **p<0.01; ***p<0.001*.

**Supplementary Figure 5.**
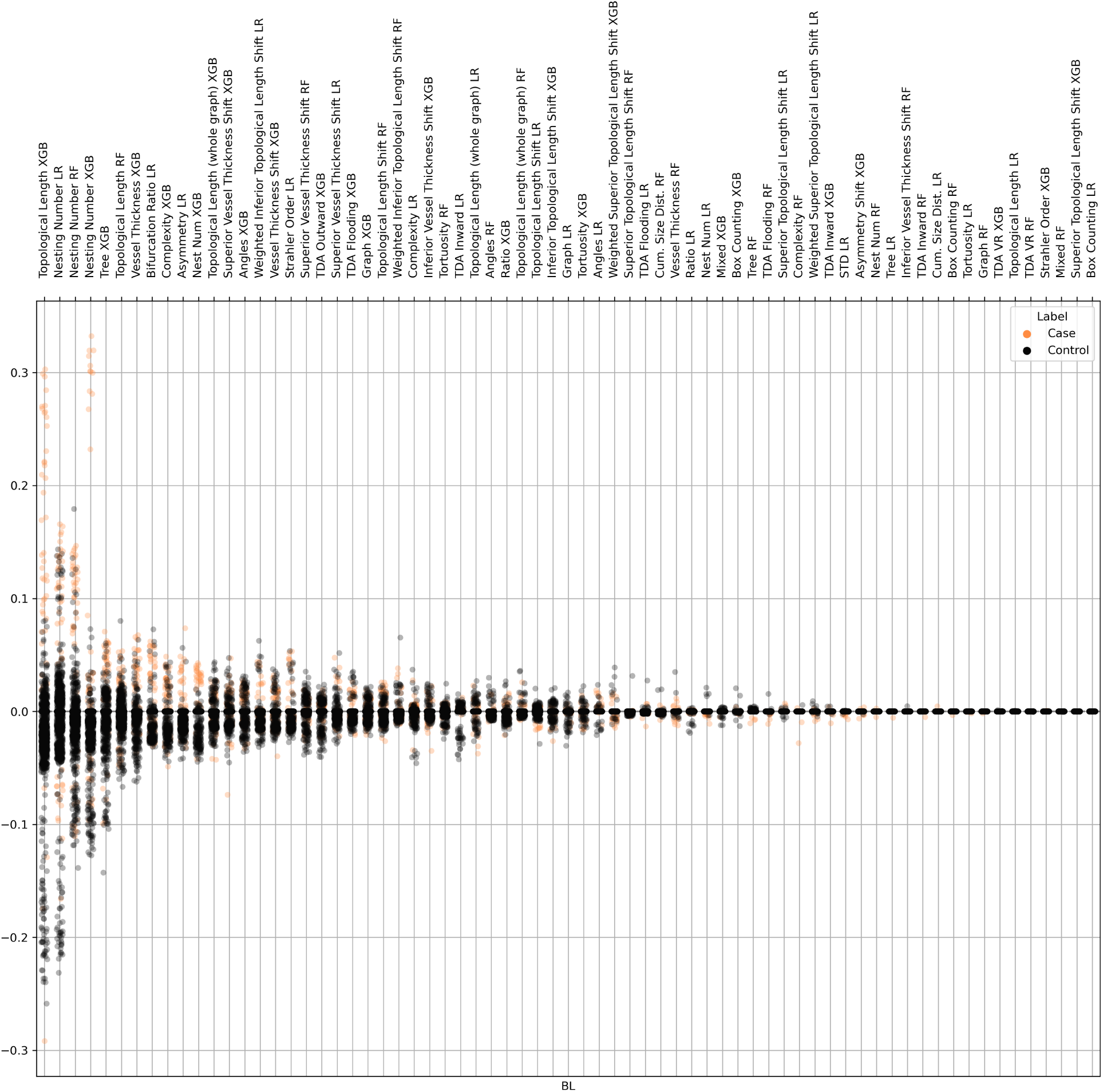
Visionary AI base learner SHAP values for predicting GHTN with Population-Wide Controls. For each population-wide model, SHAP values were calculated for each patient. These SHAP vectors were normalized and averaged by patient across models. The base learners are sorted by average absolute SHAP value.

**Supplementary Figure 6.**
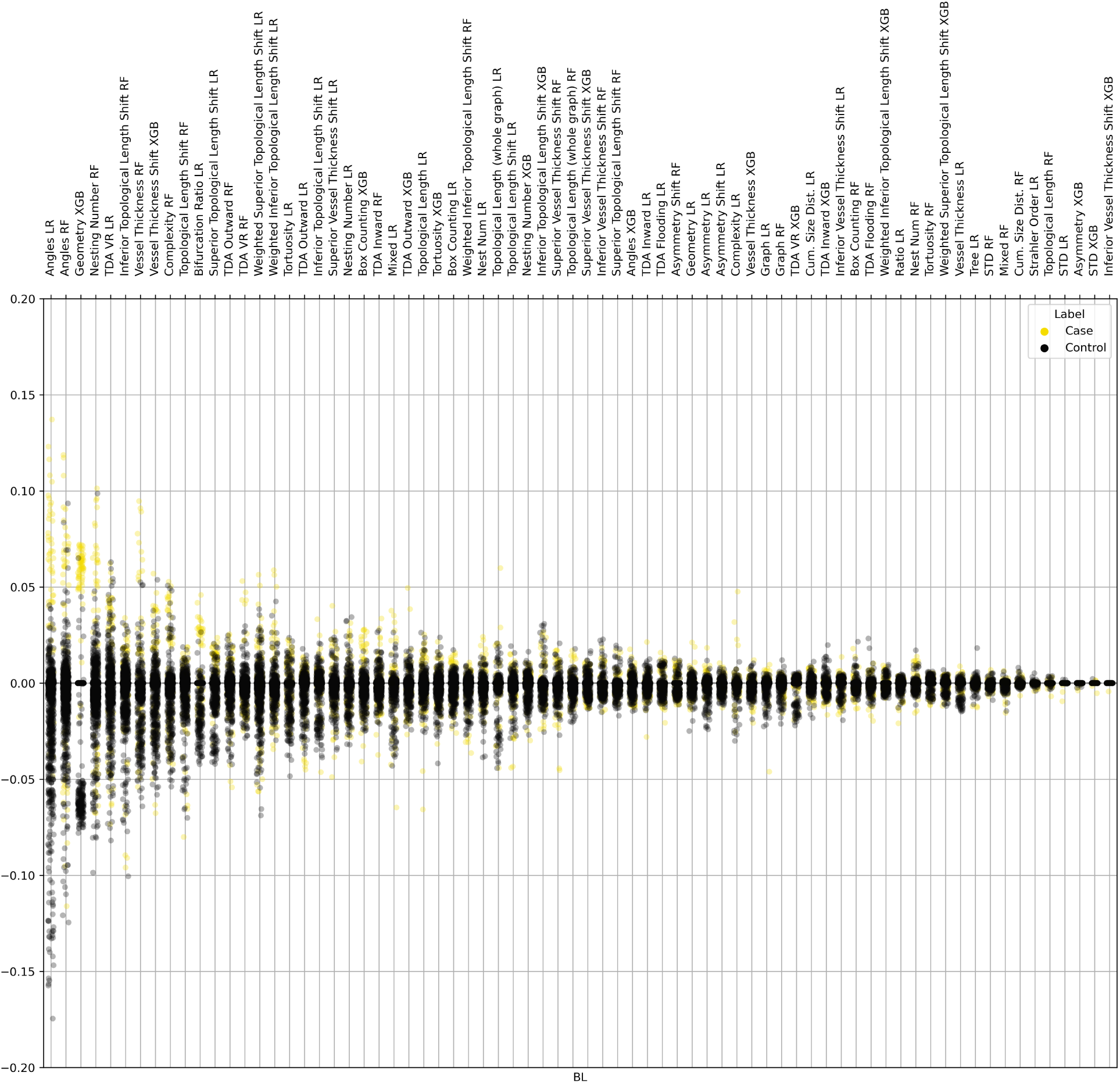
Visionary AI base learner SHAP values for predicting CHTN with Population-Wide Controls. For each population-wide model, SHAP values were calculated for each patient. These SHAP vectors were normalized and averaged by patient across models. The base learners are sorted by average absolute SHAP value.

